# Disrupted Diurnal Phase Variation in Circulating Factors as a Predictor of Disease and Mortality

**DOI:** 10.64898/2026.01.16.26344298

**Authors:** Eugene Melamud, Antonijo Mrčela, Nicholas F. Lahens, Carsten Skarke, Garret A. FitzGerald

## Abstract

Maintaining normal diurnal rhythms is critical for human health, yet due to limited longitudinal sampling, the oscillatory patterns of most circulating molecular factors and their alteration in pathology remain poorly characterized. Here, we used large-scale cross-sectional UK Biobank plasma data to identify daytime (diurnal phase) oscillations in more than 3,200 biomarkers. Most plasma components exhibited significant oscillatory patterns, including 98% of lipoproteins, 90% of complete blood count measures, and 56% of proteins. We further validated the enrichment of proteomic oscillations in an independent 48-hour serial sampling cohort. Approximately 25% of oscillatory biomarkers displayed sex-specific patterns, particularly hormones, phosphate regulators, immune mediators, and muscle proteins. Remodeling of oscillatory patterns was associated with pathological states, with many alterations detectable years before clinical diagnosis. We derived a composite time-of-day deviation risk score (TOD-RS) strongly associated with all-cause mortality (HR=1.40 per SD, *p* < 1 × 10^−16^). These findings highlight the pervasive nature of daytime oscillatory regulation, its disruption in disease, and the potential use of oscillatory biomarkers for risk stratification and early disease detection.

## Introduction

Maintenance of circadian rhythms is essential for normal homeostasis and long-term health[1–3]. Conversely, loss of these rhythms has been linked to the development of multiple diseases[4–7]. However, because large-scale diurnal biomarker sampling is scarce in prospective cohorts, the health consequences of disrupted oscillatory patterns remain poorly understood[8].

The UK Biobank (UKBB) collected blood samples from nearly 500,000 participants at baseline (2006–2010), with most samples taken between 9am and 8pm[9]. These samples were used to measure a wide range of biomarkers, including clinical chemistry measures, lipids, and plasma proteins (Fig.1a). Although these cross-sectional data do not permit characterization of full 24-hour circadian rhythms, the combination of large sample size, minute-resolution timestamps, and broad molecular profiling enables detection of systematic changes in biomarker levels across the 11-hour daytime collection window. To clarify, we refer to this daytime-restricted variation as “diurnal phase” variation, to distinguish it from full diurnal or circadian rhythms (see Terminology).

**Figure 1:**
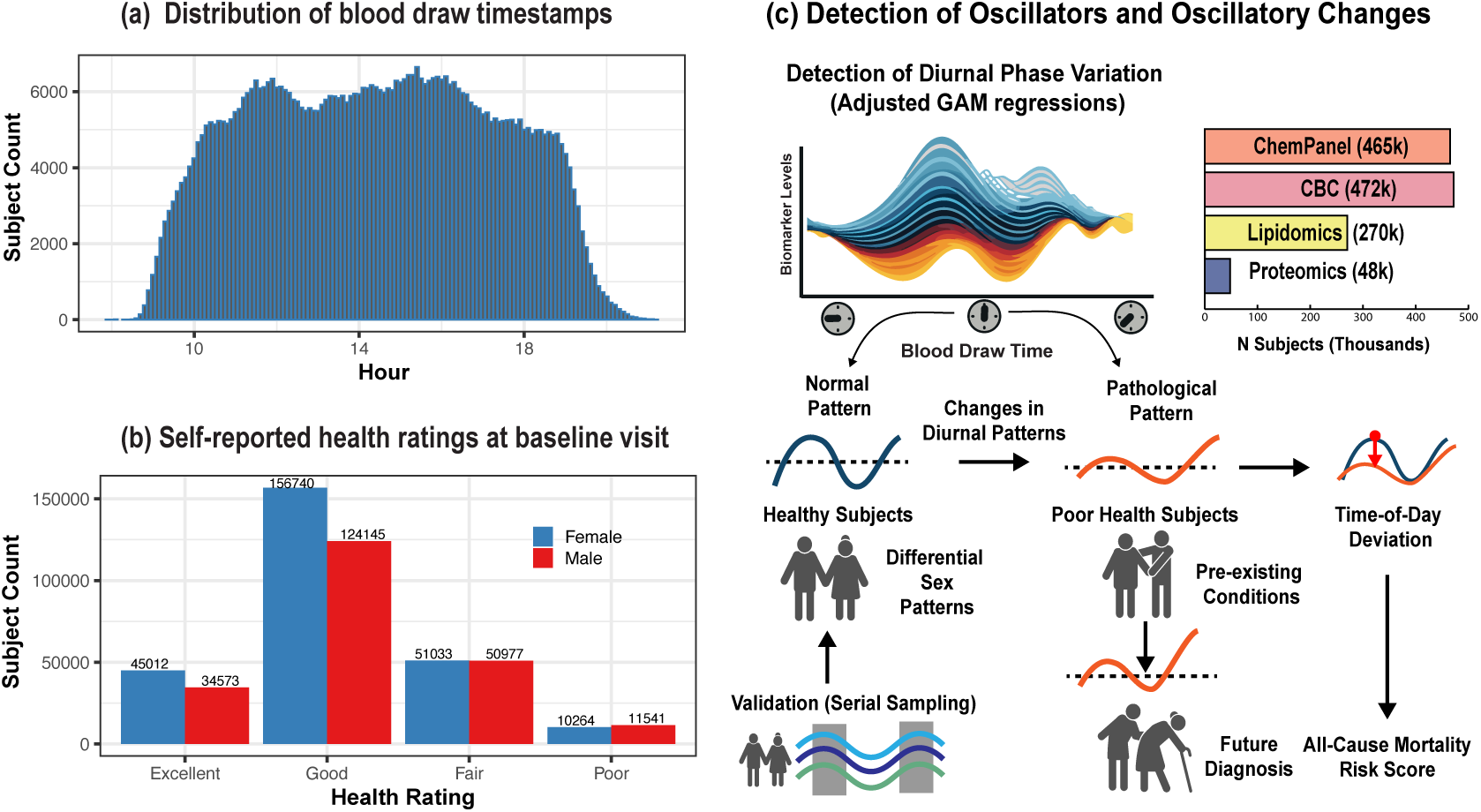
Study overview. **a**, Distribution of blood sample collection times in UKBB across all subjects at the baseline visit. **b**, Distribution of self-reported health status at baseline. **c**, To identify oscillatory biomarkers and changes in patterns of oscillations, General additive model (GAM) is used to fit day-time variation of ∼3,200 clinical and omics biomarkers. Simulations and serial sampled dataset is used to validate the findings. To identify differential oscillatory patterns linked with health, sex, pathologies, and future outcomes, the population is stratified into subgroups and differences in spline shapes are used to quantify separation in the pattern. Time-of-day deviations was used to construct risk scores and identify high-risk populations.

If oscillatory biomarkers are detectable in cross-sectional data, they would complement existing serial biomarker studies and facilitate novel oscillator discovery[10–15]. Moreover, health-related information about preexisting conditions and future outcomes is available for all subjects, enabling the assessment of changes in chronobiology with aging and the development of pathological traits. This information is particularly valuable, as blood draws were typically performed years before disease diagnosis, allowing for the testing of the hypothesis that deviations in normal oscillatory patterns preceded the development of these disorders[16–19].

Detecting oscillatory patterns in cross-sectional data poses several challenges: signal loss from insufficient sampling, variability in participants’ genetic backgrounds and environmental exposures, and biases in cohort age distribution and health status[20]. To address these issues, we applied penalized spline (GAM) regression and conducted simulations to test whether true oscillators could be recovered under realistic noise and covariance structures. We then validated the results by comparing them with oscillators identified in a 48-hour serial sampling study established oscillatory biomarkers[21]. This integrative, multi-pronged strategy enhances our confidence in the identified oscillatory biomarkers.

Our analysis shows that the plasma composition exhibits consistent diurnal variation, confirming known factors and identifying hundreds of novel oscillators. We find that healthy men and women exhibit similar oscillatory patterns and highlight sex-specific effects; these patterns are altered in the presence of pathological conditions, and the assessment of time-of-day deviation can be used for the calculation of the risk of future outcomes, such as all-cause mortality.

## Results

### Detection of Oscillators in Cross-Sectional Data

As a source of plasma biomarkers, we utilized profiling data collected at the baseline visit of the UKBB[9]. This included a 30-biomarker clinical chemistry panel and 31 CBC measures (n∼472,000), 246 lipidomic measures (n∼270,000), and 2,919 plasma proteins (n∼48,000)[22–24]. The UKBB cohort is largely representative of the general population, although with a bias towards a healthier subpopulation[25]; thus, our ability to detect normal oscillatory patterns, as well as deviations from the norms, is subject to the health characteristics of the sampled population (Fig.1b). To identify healthy oscillatory patterns, we focused our analysis on participants who self-reported being in good or excellent health (n∼360,000). In contrast, for the discovery of pathological patterns, we performed the analysis in the poor- or fair-health subset of the population (n∼123,000). For the detection of sex-specific health effects, we performed a sex-stratified analysis across all health groups.

To assess the temporal dependence of a circulating factor, we performed penalized smoothing spline regression (P-spline with 24 knots) on biomarker levels as a function of blood draw time (Fig.1c). Regressions were adjusted for age, sex, BMI, smoking status, fasting time, and sample storage time (see Methods). For non-oscillatory biomarkers, a smoothing spline fit is expected to yield an approximately flat line (null hypothesis), conversely, oscillatory biomarkers will be associated with non-linear temporal deviations (alternative model), thus providing higher model exploratory power (larger F-statistic) [26].

To test the power of the methodology to detect true oscillatory biomarkers, we conducted two types of simulations. In the first simulations, we permuted timestamps of blood draws to generate a random expectation of the spline fit statistic. This allows us to estimate the false discovery rate (FDR), while maintaining realistic covariance structures and biomarker measurement noise (Fig.2a). In the second simulation, we simulated true positive oscillatory biomarkers by generating biomarkers with random combinations of amplitude and phase shifts, alongside true negative biomarkers with zero amplitude and phase shifts (Fig.2b). Again, to maintain realism, we sample from the observed biomarker variances.

**Figure 2:**
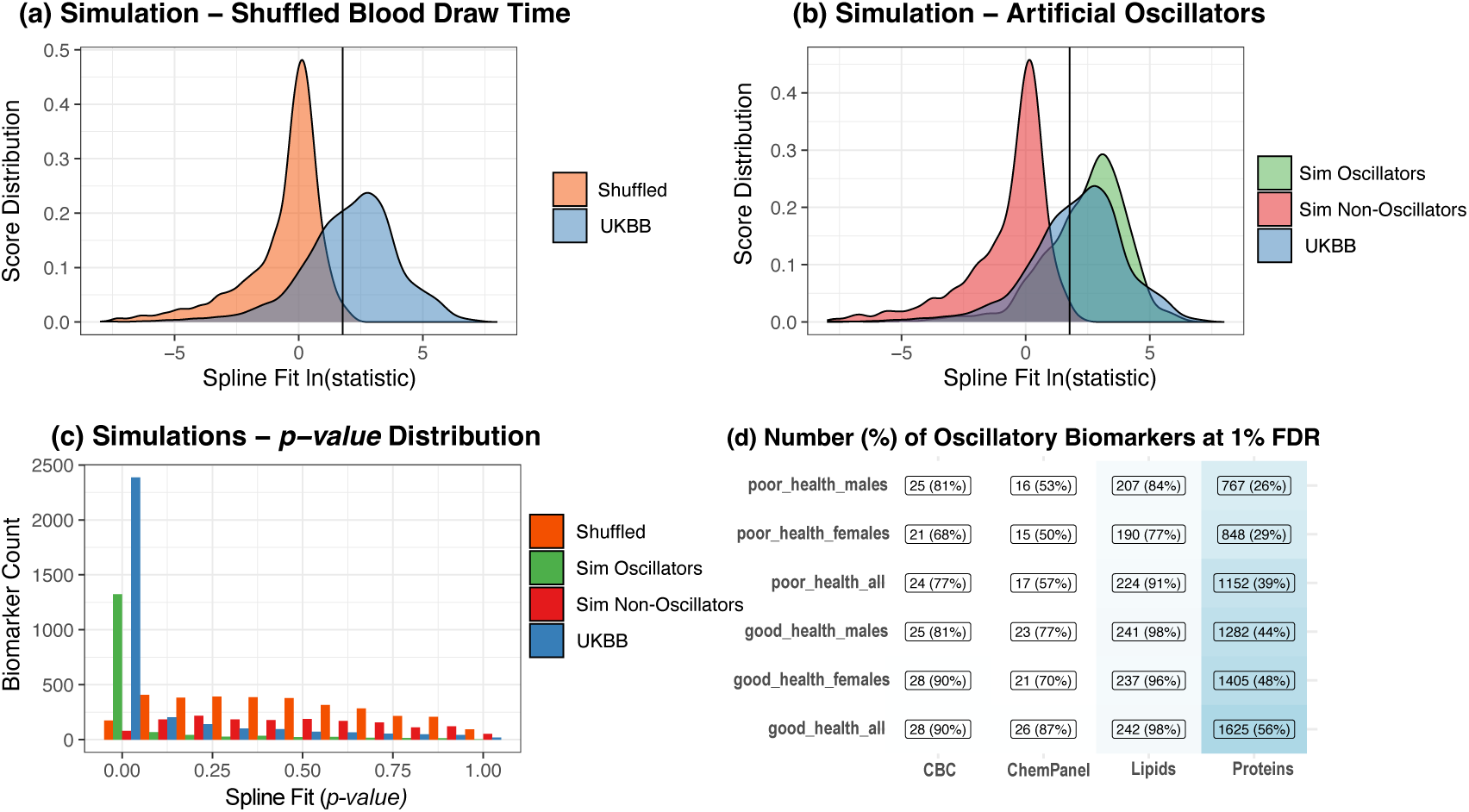
Estimated Fraction of Oscillatory Biomarkers. To estimate fraction of oscillatory biomarkers in UKBB, we detect time-of-day variation on biomarker levels using GAM spline regression, and estimate 1% false discovery rate (vertical lines) using two types of simulations. **a**, Distribution of spline F-statistic in observed vs randomly shuffled timestamp biomarker data. **b**, Distribution of spline F-statistic for artificially simulated oscillatory and non-oscillatory biomarkers versus observed biomarkers. **c**, Histogram of the *p*-values for time-of-day spline term across all simulations.

In both simulations, we find a strong enrichment in oscillatory biomarkers. The *p*-value histogram of spline fit term in simulated and observational UKBB biomarker data is shown in Fig.2c. We find that above the threshold of the spline F-statistic ⩾ 6, the false discovery rate (FDR) for oscillatory biomarkers was less than 1%. Indicating that the methodology is robust to the noise and variability present in the UKBB dataset, and hundreds of oscillatory biomarkers can be detected with high confidence.

In the healthy subset of the cohort (both men and women), 90% of CBC, 87% of the clinical chemistry panel, 98% of lipids, and 56% of proteins exhibited oscillations (Fig.2d). The fraction of oscillatory biomarkers was similar between men and women. The fraction of oscillatory biomarkers in sick individuals was significantly lower (77% of CBC, 57% chem. panel, 91% lipids, 39% of proteins). The reduced fraction among the sick is partly due to lower statistical power (fewer sick subjects) but could also reflect a genuine loss of oscillatory patterns in pathological states. We explore this topic further in the last section of the article. Complete tables of oscillatory biomarkers in healthy and sick subpopulations are provided in Tables S1 and S2.

### Comparison of Cross-sectional and Serial Oscillators

To validate further that our statistical approach can detect true oscillators, we compared oscillatory proteomics profiles from healthy subjects in UKBB (n=34,843) to the serial sampling dataset in the Chronobiome study[21]. The serial data consisted of 9 serial proteomics measurements over 48 hours in 20 healthy volunteers using the same Olin Explorer platform. Although the size of the serial dataset was much smaller, we expect an enrichment of oscillatory proteins when comparing both datasets, as well as similarity in the shape of the oscillation over the sampled time.

A plot of the 48-hour cosinor amplitude versus the spline F-statistic is shown in Fig.3a. At the 1% FDR threshold in both studies, strong concordance was observed in oscillatory proteins. Specifically, concordance was observed for approximately 66% of the proteins analyzed (1004 oscillatory proteins and 930 non-oscillatory proteins). However, we also identified instances of discordance: 327 oscillatory proteins were detected only in the chronobiome study, while 645 were detected only in the present study (Table S3). These discordances can be attributed to differences in statistical power between the two studies, variations in the monitoring period (12 hours vs. 48 hours), and differences in the health characteristics of the subjects. Despite these discrepancies, our results indicate a strong enrichment of serial oscillators within the UKBB dataset (*p* < 2.2 × 10^−16^ Fisher Exact Test).

**Figure 3:**
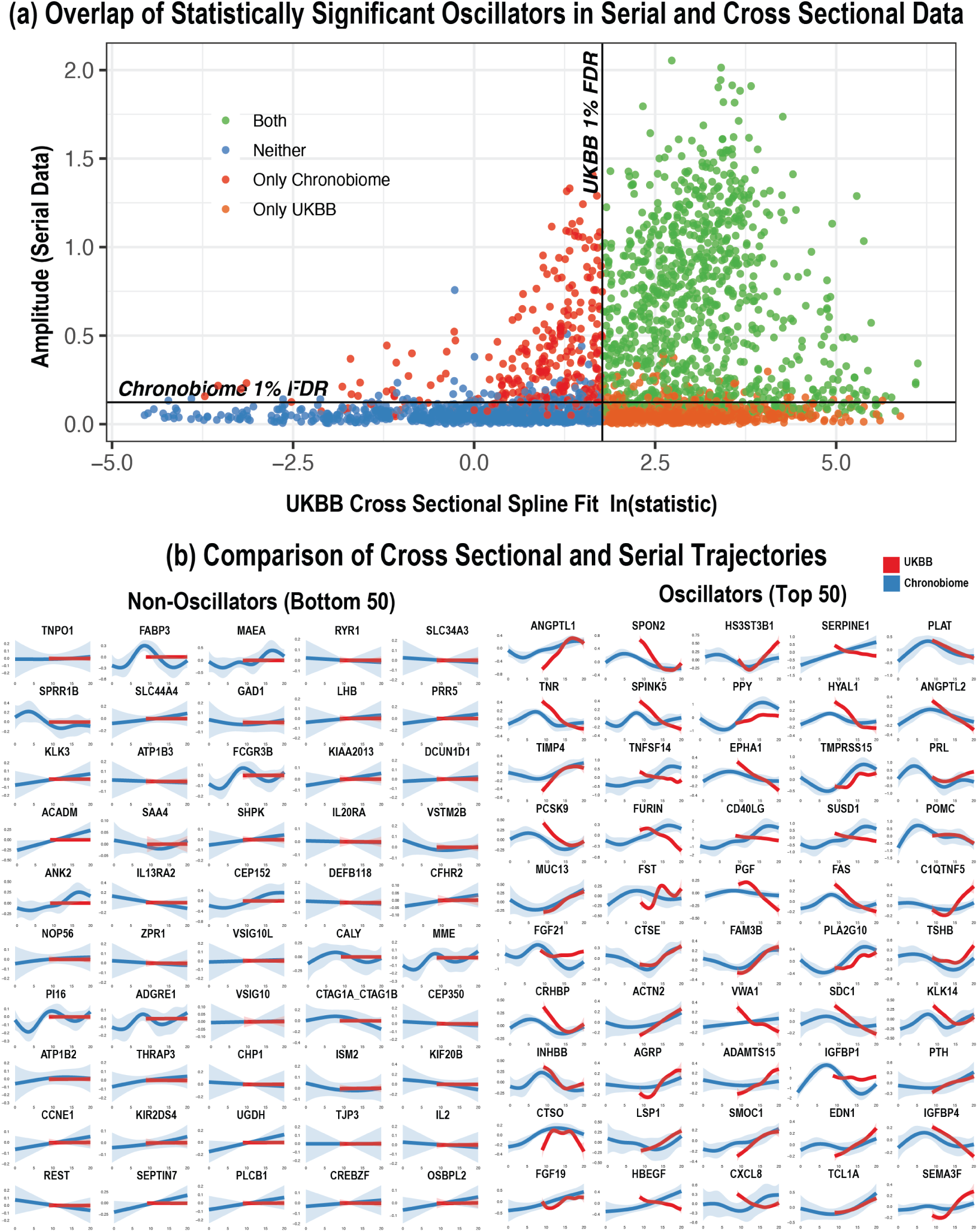
Comparison of cross-sectionaly and serially derived oscillators. **a**, Comparison of significant proteomics oscillators identified in healthy subjects in UKBB and the serially sampled Chronobiome dataset. Horizontal line represents 1% FDR spline F-statistic in UKBB. Vertical line represents 1% FDR cosinor fit in Chronobiome. Strong concordance (66%) of non-oscillatory and oscillatory proteinswas observed between the two datasets. **b**, Comparison of spline shape for top 50 UKBB oscillatory and non-oscillatory proteins in healthy subjects (11 hours, n=34,843) vs. spline fit in serial data (24 hours, n=20).

We further validated the concordance between the two datasets by comparing the shapes of the fitted splines(see Methods). Correlations were computed using the Pearson correlation coefficient over the shared daytime window. The strongest UKBB oscillators showed the highest concordance (average *r* > 0.5 at ln(F-statistic)> 5), with average correlations declining for weaker oscillators (Fig.S4). A visualization of spline fits for the top 50 UKBB oscillators and non-oscillators is shown in Fig.3b. Although the shapes of the splines were not identical, similar patterns were observed for most proteins, indicating that the cross-sectional spline approach can be used to approximate real oscillatory behavior.

### Normal Oscillatory Patterns in Healthy Subjects

To explore the biology of diurnal phase biomarkers, we focused on those with the largest oscillatory amplitude (peak–trough within the observed window). The oscillatory amplitude (Fig.4a) and spline shapes (Fig.4b) were similar in men and women (*r_p_* = 0.89, *p* < 2.2 × 10^−16^). The oscillatory patterns for the top 50 oscillators in each biomarker category are shown as heatmaps in Fig.4c.

**Figure 4:**
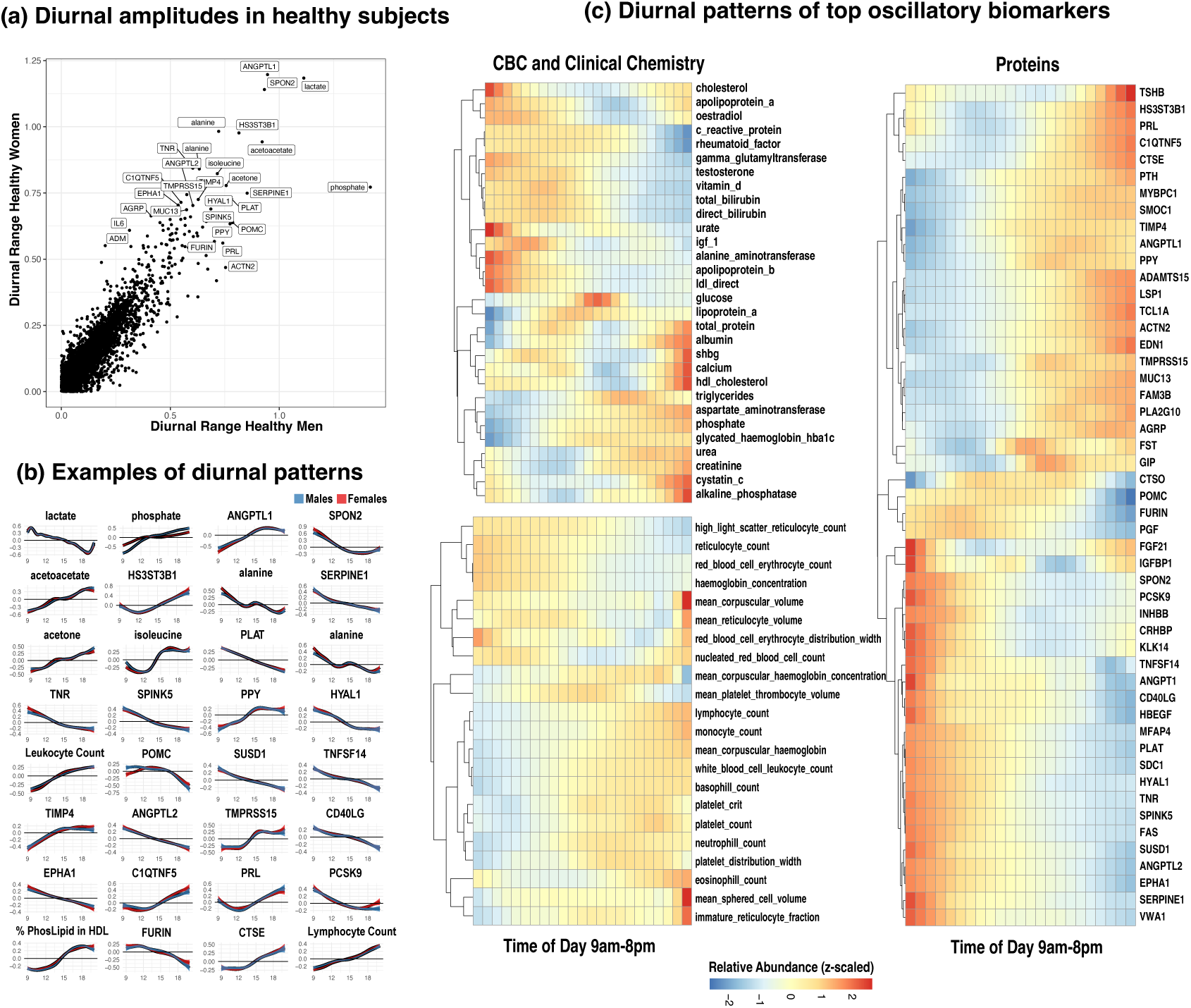
Diurnal phase patterns in healthy individuals. **a**, Comparison of oscillatory amplitudes in healthy male and female subjects. **b**, Examples of biomarkers with largest oscillatory amplitude. Overall, men and women show similar patterns of daytime variation. **c**, Heatmap of CBC, clinical chemistry, and top 50 protein oscillatory biomarkers with the largest oscillatory amplitude plotted in half-hour intervals.

Plasma metabolite profiles displayed significant variation. Glycolysis/gluconeogenesis products, such as lactate, alanine, and pyruvate, decreased over the course of the daytime hours. In contrast, ketogenesis-related metabolites (acetone, acetoacetate, and hydroxybutyrate) increased throughout the day (Fig.S6). Furthermore, concentrations of branched-chain amino acids peaked at 3pm, which may be attributable to patterns of protein-rich dietary intake in the afternoon. Glucose levels also showed an increase in the afternoon in women but not in men. Circulating triglyceride and fatty acids concentrations reached their maximum values in the evening, potentially indicating the intake of fatty meals.

More than a thousand proteins exhibited significant oscillations. Among the strongest oscillators were a wide range of growth factors (PGF, FGF21, FGF19, PDGFB, PDGFA, EGF) as well as hormones and tissue maintenance proteins (POMC, PRL, PTH, EDN1, EPO, GCG, GRP, GIP, GDF15, TSHB, PPY). We also observed significant changes in cytokines and regulators of immune cell activity (SPON2, IL6, LGALS1, TNFSF14, CD40LG, CCL15, NECTIN2). Heatmaps of the top proteomics oscillators are shown in Fig.4c. The most prominent changes were observed in Angiopoietin-like proteins, with an increase in ANGPTL1 levels and a decrease in ANGPTL2 levels. Both proteins play important roles in metabolism and tissue repair and maintenance processes[27]. We also observed changes in both the diameter and composition of cholesterol and phospholipids in LDL/HDL particles (a decrease in cholesterol in LDL and an increase in cholesterol in HDL). Interestingly, plasma levels of PCSK9 decreased throughout the day, which could indicate differential regulation of LDL receptors in the liver and the regulation of lipoproteins. Notably, similar observations of diurnal regulation of LDL by PCSK9 have been made with implications of optimal effectiveness of time-of-day specific treatment of PCSK9 inhibitors[28, 29].

The blood cell compositions in plasma also exhibited significant variation. All white blood cell counts (lymphocytes, neutrophils, eosinophils, basophils) and platelet counts increased throughout the day (Fig.S5a). In contrast, red blood cell counts, reticulocyte counts, and hemoglobin concentrations decreased. The temporal variation in white blood cells and platelets indicates that the immune system and tissue repair processes are strongly time-dependent[30].

Among ions, phosphate levels exhibited the largest oscillatory amplitude, rising throughout the sampled interval. These findings were consistent with previous reports of circadian rhythms in phosphate metabolism[31–33]. The increase in phosphate was more pronounced in men compared to women, suggesting possible sex-specific hormonal regulation of phosphate metabolism. To investigate this further, we examined whether known regulators of phosphate exhibit oscillatory patterns (Fig.S7). We observed significant daytime variation in PTH and EPO, along with small sex-specific oscillatory changes in the levels of FGF23. The direction of the regulatory effect is challenging to ascertain, as it is possible that the increase in PTH and EPO is a response to the rise in phosphate levels. Other regulators of phosphate levels and biomarkers of kidney function remained largely unchanged.

### Oscillatory Differences Between Sexes

Normal, sex-related differences in circadian regulation are expected to influence biomarker variation[34]. We therefore asked whether sex-specific differences in oscillations could be identified in the healthy subpopulation. We computed covariate-adjusted spline fits separately in healthy men (n∼158,720) and women (n∼201,750), subtracted the fitted spline shapes, and assessed whether there were significant differences in spline shape and oscillatory amplitude between groups (see Methods). A plot of sex-specific oscillatory amplitude versus the magnitude of spline separation is shown in Fig.5. At 1% FDR, we detected 740 (25%) sex-differential oscillatory biomarkers (Table S4).

**Figure 5:**
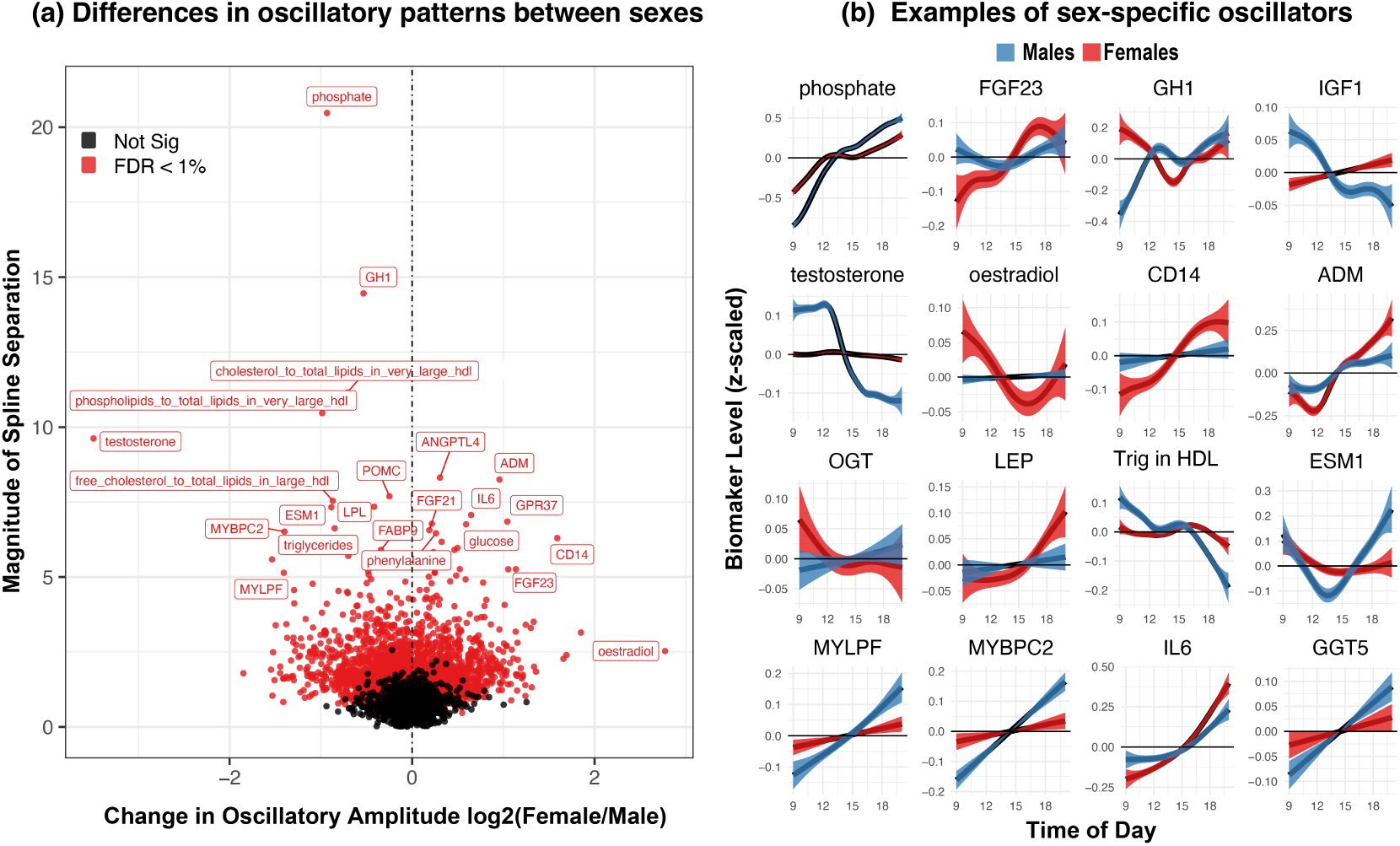
Sex-differential oscillators. **a**, Log2 fold difference in oscillatory amplitude of healthy females vs. males. The y-axis represents difference is spline shape as defined by magnitude of spline separation. **b**, Examples of biomarkers with the largest sex-differential effects.

To assess robustness, we repeated the detection of oscillatory biomarkers in a smaller subset of healthy subjects from the second UKBB visit (n∼15,700). The analysis was limited to 317 non-proteomic biomarkers measured at both visits. A comparison of oscillatory amplitudes between baseline and the second visit is shown in Fig.S3. Oscillatory amplitudes were highly reproducible, with similar ranges and patterns in both sexes (*r_p_* = 0.83, *p* < 2.2 × 10−16).

The largest differences were observed in the sex hormones testosterone and estradiol. Levels of both hormones were higher in the morning and declined rapidly by the afternoon. The daytime decline in sex hormones has been described in previous studies[35–37], although we note that the sharp transition in testosterone levels was notably different in this study. Diurnal variation in Growth Hormone (GH1) has also been previously described[31, 38]. We observed a steady increase in GH1 throughout the day in men, whereas women exhibited a dipping pattern toward the afternoon. As noted previously, we also observed significant sex-specific differences in the levels of phosphate and FGF23 between the sexes, with a sharper rise in phosphate and a shallower increase in FGF23 in men compared to women.

We also observed sex-specific differences in the oscillations of skeletal muscle proteins MYBPC2 (Myosin Binding Protein C, Fast Type) and MYLPF (Myosin Light Chain, Phosphoryl table, Fast Skeletal Muscle). Both proteins are involved in the processes of skeletal muscle contraction; the increase in circulating levels of these proteins may indicate higher muscle turnover or usage in males. Other notable sex-specific differences were observed in immune system proteins, including chemokine CCL15, pattern recognition receptor CD14, IL-6 cytokine, and Endocan (ESM1), which modulates the adhesion of leukocytes to endothelial cells. Although the sources of these proteins in plasma could originate from a diverse set of tissues and cell types, it is plausible that their variations are driven by differential immune system responses to environmental exposures.

### Oscillatory Changes in Pathological Conditions

The global effects of pathology could manifest as changes in both plasma levels and the pattern of oscillations in circulating factors (Fig.S8). To evaluate the effect of pathologies on biomarker oscillations (independently of changes in biomarker levels), we carried out a number of comparisons between sick versus healthy subgroups (self-reported health), subjects with pre-existing conditions, and post baseline future diagnoses(ICD-10 level codes). Following the same procedure as described in the previous section, we stratified the population into subgroups, calculated adjusted spline fits, and compared the differences in the magnitude of spline separation between sub cohorts.

Comparing sick and healthy subgroups, at 1%FDR, approximately 15% of biomarkers (n=492) showed a significant difference in oscillatory patterns (Fig.6a). The largest differences were observed in biomarkers linked with metabolic disorders, such as glucose, fatty-acid binding proteins (FABPB1, FABP3), Glutathione S-transferases (GSTA1, GSTA3), and Sulfotransferase Family 2A Member 1 (SULT2A1). This observation was consistent, as metabolic disorders are strongly enriched among the sick (Fig.S10).

To gain further specificity, we examined differences in oscillatory patterns at the ICD-10 code level. The most common conditions enriched in the sick subjects included hypertension (I10), hyperlipidemias (E78), joint disorders (M25), asthma (J45), cardiovascular diseases (I20, I21), and diabetes (E14 and E11). Although our power to detect changes in specific diseases was reduced (fewer diagnosed cases), we observed that a large fraction of biomarkers is affected by pre-existing conditions (Fig.S10b, Table S5 and S6). A similar pattern was observed when stratifying subjects into groups with yes/no future ICD-10 code diagnoses. We found a substantial fraction of biomarkers (5-15%) already had a differential oscillatory pattern at baseline, suggesting that these biomarker changes potentially preceded disease diagnosis. Although caution is warranted, it is possible that many subjects may have had undiagnosed and comorbid conditions at baseline.

Multiple biomarkers, including glucose, CFD, ANGPTL4, FABP1, and FST—were associated with multiple diseases, likely reflecting metabolic syndrome comorbidity[39–41]. We therefore examined oscillatory changes specifically in type 2 diabetes (E11). Approximately 25% of biomarkers showed loss or distortion of their normal oscillatory pattern. Many biomarkers that exhibited the largest oscillatory changes in diagnosed diabetes also displayed substantial changes in individuals who were diabetes-free at baseline but later received a diagnosis, indicating that altered oscillatory patterns are already present prior to clinical recognition (Figure 6c). The most pronounced difference was in glucose, with higher morning peaks that declined toward the afternoon; as expected, absolute glucose levels were elevated across the sampled window in diabetes[42], demonstrating disruption in both level and pattern (Figure 6d).

**Figure 6:**
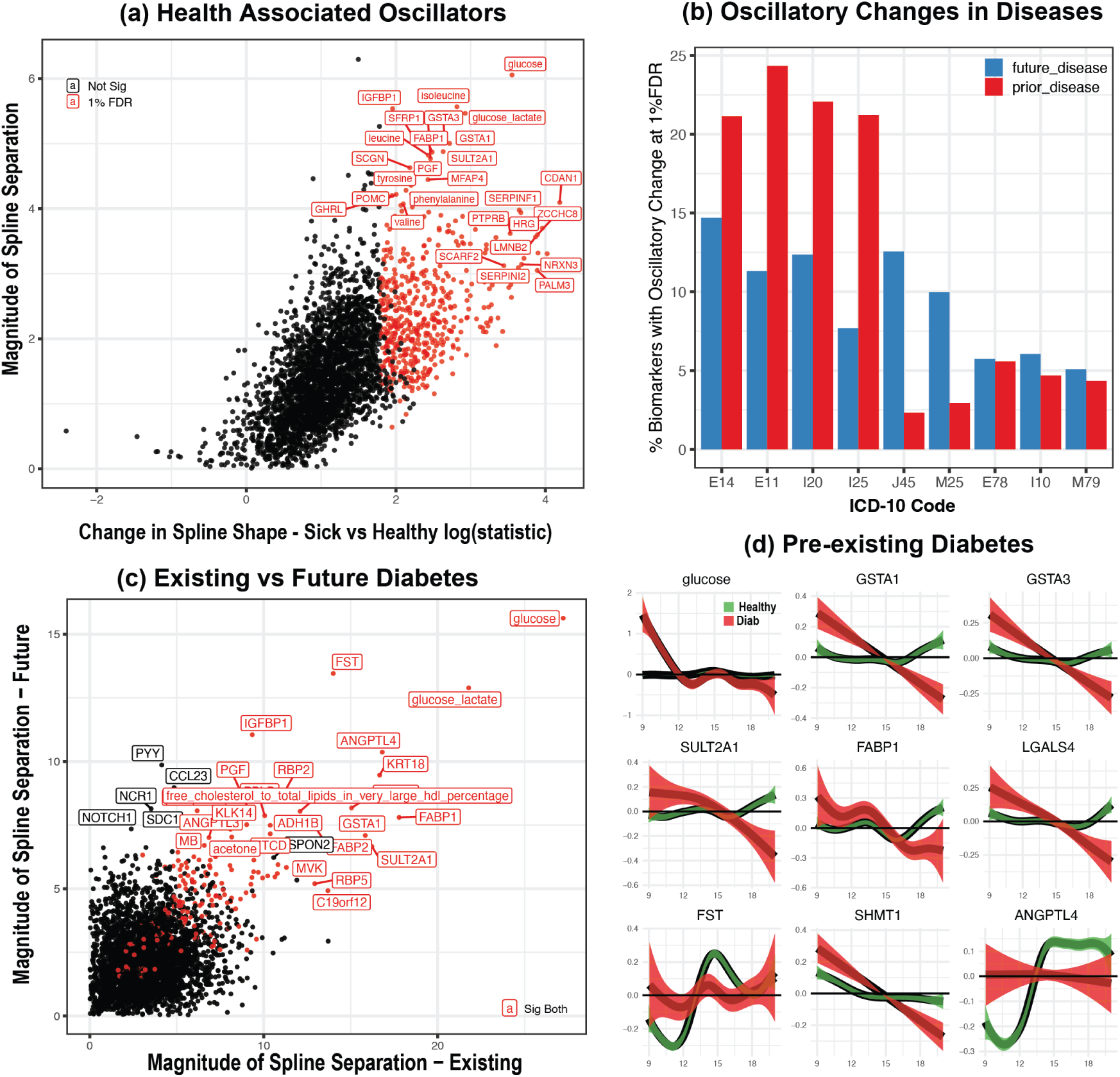
Oscillatory differences between sick and healthy. **a**, Statistically significant changes between sick and healthy. **b**, Fraction of oscillatory changes associated with specific ICD10 diagnostic code. Comparison of subjects with pre-existing or future diagnosis vs healthy subcohort. **c,d**, Comparison of oscillatory changes in subjects who will be diagnosed with diabetes in the future (2+ years) vs no diagnosis. Spline separation defined as absolute sum of difference between spline shapes.

Overall, oscillatory patterns are substantially altered in pathological conditions and are associated with future adverse outcomes. However, beyond a few common diseases, the low number of diagnosed cases and limited biomarker coverage (particularly for proteomics) constrain our power to detect disease-associated changes; thus, we are likely underestimating the true extent of alterations in oscillatory patterns in pathological states.

### Oscillatory Changes and All-Cause Mortality

We next evaluated associations between oscillatory patterns and all-cause mortality. Most deaths arise in the context of pre-existing disease and multimorbidity, and circulating biomarker levels are known predictors of adverse outcomes[43]. Whether mortality risk is also accompanied by altered oscillatory patterns is less clear.

We compared covariate-adjusted spline fits between participants who died and those who survived over 16 years of follow-up, quantified the magnitude of spline separation, and estimated biomarker level associations with all-cause mortality using Cox proportional hazards (CoxPH) regression (see Methods).

The relationship between ln(HR) for biomarker levels and the magnitude of spline separation is shown in Fig.7a.

**Figure 7:**
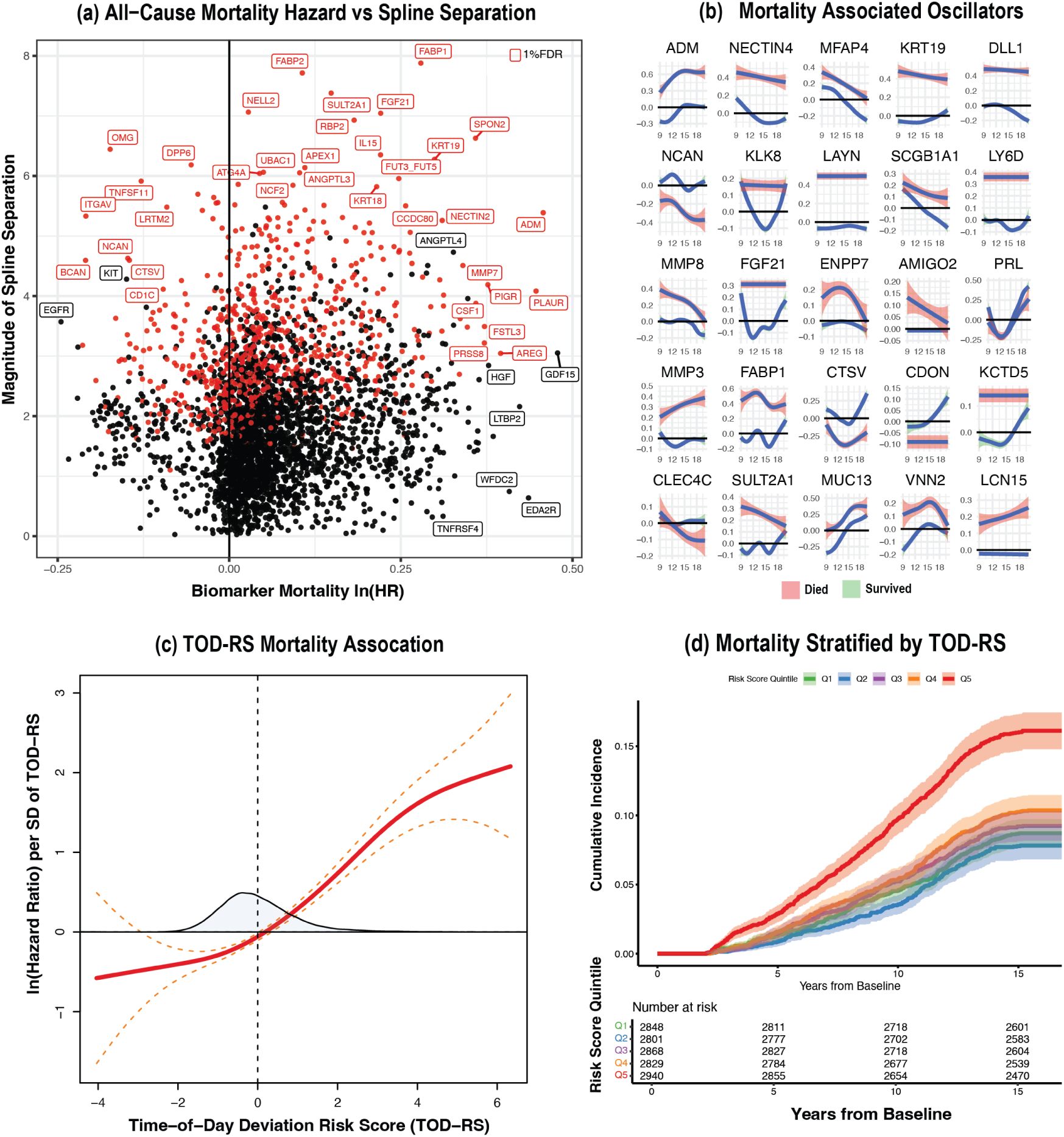
Association of changes in oscillatory patterns with all-cause mortality. **a**, Biomarker association with all-cause mortality (CoxPH ln(HR)) vs. magnitude of spline separation in the biomarker between those that died and survived. Biomarkers with significant change in oscillatory pattern (1% FDR) are shown in red. **b**, Examples of mortality associated oscillators. **c**, Association of time-of-day deviation risk score (TOD-RS) with all cause mortality (pspline CoxPH on test subset). **d**, KM Plot of stratified on TOD-RS with all-cause-mortality.

At 1% FDR, 22% of biomarkers (n=751) exhibited significantly different oscillatory patterns between participants who died and those who survived (Table S7). Of these, 71% (n=524) also showed level associations with all-cause mortality. Protein biomarkers with a pronounced oscillatory pattern disruption included lipid metabolism factors (FABP1, FABP2, ENPP7, PCSK9), hormones (ADM, FGF21, PRL), immune and inflammatory mediators (TNFSF11, IDO1, IL15, IL32), matrix remodeling enzymes (MMP8, MMP3, MMP7), and cell adhesion molecules (NECTIN4, NECTIN2, NCAN, DLL1, KCTD5, ITGAV).

Based on these observations, we sought to derive a composite score of oscillatory pattern disruption and its association with all-cause mortality. We selected the top 100 mortality-associated oscillators, defined as those with the largest magnitude of spline separation and confirmed as oscillatory in the Chronobiome study (passing 1% FDR in both studies). For all subjects, we computed each individual’s time-of-day deviation from the expected biomarker level for a healthy subject at the same sampling time (see Methods). To prune redundant signals, we used gradient boosted models (GBMs) to select oscillators with the strongest association with mortality (Table S8, Fig.S11)[44]. The model was trained on 70% (n=28,387) of subjects with proteomics data and evaluated on the remaining 30% (n=12,196). Twenty-six oscillators were selected by GBMs and used to compute the Time-of-Day Deviation Risk Score (TOD-RS) for each subject((Fig.7b, Fig.S11e-f).

The Time-of-Day Deviation Risk Score (TOD-RS) captures the aggregate disruption of daytime oscillatory patterns across selected biomarkers and is significantly associated with all-cause mortality in both training (CoxPH HR=1.46 per SD TOD-RS, *p* < 2.2 × 10^−16^) and test subsets (HR=1.40 per SD, *p* < 2.2 × 10−^16^), indicating that greater disruption is linked to a higher risk. For visualization, we plotted the hazard as a continuous function of TOD-RS using a penalized spline term (Fig.7c) and stratified subjects by TOD-RS quintiles in the Kaplan–Meier survival analysis (Fig.7d). The highest quintile showed a 2.37-fold higher all-cause mortality relative to the lowest (HR=2.37, *p* < 2.2 × 10^−16^).

Notably, although TOD-RS–selected biomarkers are enriched for oscillators with the largest deviations in their oscillatory range, they are not necessarily enriched for biomarkers with the highest mortality hazard. For example, biomarkers such as GDF15, WFDC2, and Cystatin C are among the strongest mortality-associated biomarkers by level; however, they do not show a significant difference in oscillatory patterns between those who died and those who survived. Thus, TOD-RS captures only a fraction of biomarker risk limited to oscillators rather than the full set of biomarkers.

## Discussion

Circadian variation in plasma composition has been extensively investigated, with most previous investigations relying on serial sampling of a small number of subjects to discover oscillatory biomarkers[11–13, 15, 45–49]. In this study, we leverage the scale of UKBB data to describe diurnal phase variation in major components of plasma composition under both normal and pathological conditions. Despite the challenges posed by cross-sectional data, we demonstrate through theoretical simulations and comparisons with serially collected data from the human Chronobiome study that the spline fitting approach can recover high-confidence oscillators[21].

We demonstrate that, in healthy subjects, over half of proteins and nearly all blood cell counts, lipoprotein levels, and metabolites exhibit a measurable oscillatory pattern. Although the large effects of circadian rhythm on plasma composition align with previous serial sampling studies[10, 16, 47, 49, 50], the numerous oscillators in this study likely result from the increased power of the UKBB cohort. Our expectation is that as the scale of sampling in human cohorts increases, an even greater fraction of biomarkers will be found to exhibit oscillatory patterns.

Overall, healthy men and women exhibit similar oscillatory patterns, with about a quarter of oscillators showing differential sex-specific effects. This sex-differential effect encompasses not only sex hormones but also various biomarkers, including circulating lipids, growth hormones, immune mediators, muscle proteins, and phosphate regulators.

Pathological conditions—whether defined by self-reported poor health or ICD-10 diagnoses—were frequently accompanied by changes in daytime oscillatory patterns across many biomarkers. Most effects reflected distortion of the normal pattern rather than complete loss of oscillation, though both dampening and amplification occurred. The largest alterations were linked to metabolic disorders, especially diabetes, consistent with prior reports on diabetes–circadian interactions[45]. Importantly, many oscillatory deviations were present both in established disease and in subjects who later developed the condition, inferring that circadian deconsolidation coexists with and may precede clinical diagnosis. Although, we cannot exclude the possibility that some subjects had undiagnosed comorbid diseases at the time of sampling.

To understand the relationship between changes in oscillatory patterns and long-term survival, we examined the association between oscillators and all-cause mortality. As all-cause mortality is often driven by pre-existing conditions and multimorbidity[51], we anticipated that many biomarkers predictive of mortality would also exhibit altered diurnal phase variation[43]. Indeed, our analysis confirmed that subjects who died during the 16-year follow-up period exhibited significant alterations in oscillatory patterns. To quantify the impact of altered oscillators on mortality risk, we developed a composite Time-of-Day Deviation Risk Score (TOD-RS), and show that it was strongly associated with all-cause mortality. The TOD-RS captures a subset of mortality risk focused on oscillatory disruption, and thus provides a complementary perspective on biomarker risk assessment to traditional level-based measures.

As most plasma components show diurnal phase variation, our findings underscore the need to incorporate sampling time when interpreting clinical data; failure to do so may contribute to misclassification or misdiagnosis[52, 53]. With larger datasets, time-of-day adjusted reference intervals and models could be developed, and the degree of diurnal deviation may itself predict disease risk. Models that do not account for oscillations likely miss an important component of temporal relationships with future outcomes.

Our study has several limitations. While we observed robust replication of oscillatory biomarkers across UKBB visits and serially collected data, fundamental limitations arise when using cross-sectional data to derive oscillatory patterns. False positives may arise from unmeasured confounders, such as dietary variation, physical activity, stress, and sleep disruption. Our ability to detect pathological patterns is limited by low case numbers for many conditions. Most blood samples were collected during the day, thus we likely underestimating the true oscillatory amplitudes. Finally, our findings are based on a single cohort and require replication in more diverse ethnic groups and confirmation through larger serial sampling studies.

In summary, we catalog widespread diurnal phase variation across healthy and pathological states and show that integrating large cross-sectional spline models with serial validation recovers high-confidence oscillators. We further demonstrate that the deconsolidation of normal oscillatory patterns predicts long-term mortality outcomes, supporting the importance of incorporating time-dependent biomarker levels in risk assessments. Despite the limitations of this study, our results are consistent with the hypothesis that the deconsolidation of circadian rhythms reflects disease and predicts both its emergence and mortality. With additional serial sampling, and integration of wearables and sleep data, more personalized biomarker based models of diurnal rhythm disruption and disease risk may be achievable.

## Methods

### Terminology

For consistency, we use the following terms throughout:

- **Diurnal phase variation**: Variation in biomarker levels that occurs during the daytime hours (9am-8pm).
- **Oscillatory pattern**: Spline shape of biomarker levels over the sampled daytime window.
- **Oscillatory pattern disruption**: Departure of an individual’s biomarker level from the expected healthy oscillatory pattern at the sampled time.
- **Oscillatory biomarker**: A biomarker with a significant spline term (spline F-statistic ≥ 6.0, empirically FDR < 1%).
- **Oscillatory amplitude**: Peak–trough difference of the fitted spline within the observed window; a lower bound on full 24h amplitude.
- **Magnitude spline separation**: Sum of absolute pointwise differences between two fitted oscillatory patterns.
- **Diurnal rhythm**: A biological process that exhibits a 24-hour pattern, which may be driven by endogenous circadian rhythms and/or external factors (e.g. light-dark cycle, feeding-fasting cycle).
- **Circadian rhythms**: Diurnal oscillation driven by the molecular clock.

### Plasma Biomarkers Data

The time of blood sample collection was obtained from the UKBB field 3166 and encoded as the time of day with a minute resolution. The majority (99.9%) of visits occurred between the hours of 9:00am and 8:00pm. Visits outside this range were not used in the analysis. To facilitate comparison of oscillations between different biomarkers, all biomarkers were z-scaled (mean=0, sd=1) prior to analysis. The following categories of biomarkers were used in the analysis: CBC measures (category: 100081), established biomarkers (category: 17518), lipidomics (category: 220) and proteomics (category: 1839). Biomarkers collected at the baseline visit (instance = 0) were used for bulk of the analysis. For replication, we used biomarkers collected at the second visit (instance = 1).

Disease events were obtained from the first-occurrence category (category: 1712). We defined pre-existing conditions as disease cases that occurred prior to within two years of the baseline visit. When appropriate, regressions were adjusted for the following covariate structure: age at visit (field: 21003), sex (field: 31), town deprivation index (field: 189), smoking (field: 1239), and BMI (field: 21001). We further adjusted the regression for the fasting time (field: 74) and the sample storage time (difference in time between the initial *i* visit and last date of data collection in a cohort).

### Detection of Oscillatory Biomarkers

Daytime biomarker variation was detected using a generalized additive model (GAM)[54], fit with the mgcv R package (v.1.9-1, bam function for large data). Z-scaled biomarker values were modeled as a function of time of day using a penalized P-spline (24 knots, bs=’ps’, gamma=1):

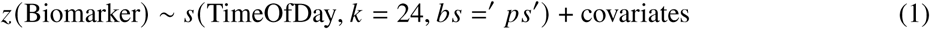

Here, *z*(Biomarker) is the standardized biomarker level and *s* (TimeOfDay, *k* = 24, *bs* =′*ps*′) is a penalized smoothing spline with 24 knots with gamma=1 to prevent overfitting.

Models were adjusted for age, sex, smoking status, Townsend deprivation index, fasting time, and sample storage time. Sex and smoking were binary; all other covariates were continuous. Spline term significance was evaluated via F-test summary statistic reported by mgcv::summary.

Oscillatory amplitude was defined as the peak–trough difference of the fitted spline (Fig.S1). Because only daytime is observed, this is a lower bound on the full 24 h amplitude.

### Detection of oscillatory pattern differences between subgroups

Detection of spline pattern changes between two subgroups (e.g. sex, health status, mortality) was performed using a two-step procedure. First, we fitted adjusted splines for each subgroup separately using the same covariate structure as in Eq.1. Second, we subtracted fitted splines to obtain the difference in the magnitude of spline separation (oscillatory pattern difference metric) between subgroups.

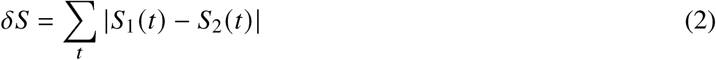

where *S*_1_(*t*) and *S*_2_(*t*) are the fitted splines for groups 1 and 2 at time *t*.

We then assess if the resulting shape is statistically different from the flat line using GAM regression. The procedure is illustrated in Fig.S1. To ensure that spline differences are robust, the procedure above was bootstrapped by repeating the spline fitting and subtraction 10 times.

### Power to detect difference in spline patterns

Power simulations were conducted to determine the sample sizes required for: (1) detecting time-of-day variation in biomarker levels and (2) detecting differences in biomarker spline shapes between groups of different sizes.

To assess the power to detect time-of-day variation under realistic conditions, we used real biomarker data and covariate structures from the top 100 oscillators in healthy subjects. To assess power, we subsampled subjects to create datasets of varying sizes (100, 500, 1,000, 2,000, 5,000, 10,000, 20,000, 40,000) and fitted a covariate-adjusted spline regression to each(Eq.1). We then determined the fraction of subsampled datasets that detected time-of-day variation at Bonferroni adjusted *p* = 3.125 × 10^−6^ as the alpha threshold, assuming 3,200 biomarker tests (0.01/3200) (Fig.S2a).

To assess the power to detect differences in spline shape between two subgroups, we simulated realistic conditions using two non-overlapping groups of subjects (diabetes vs. non-diabetes). We randomly subsampled subjects from each group to create datasets of varying sizes (50, 100, 500, 2,000, 4,000, 6,000, 10,000, 20,000). Using glucose as a known oscillatory biomarker, we assessed the fraction of simulated datasets that successfully detected a change in the oscillatory pattern(see Eq.2). We used a Bonferroni-corrected *p* = 3.125 × 10^−6^ as the alpha threshold, assuming 3,200 biomarker tests(Fig.S2b).

### Derivation of Time-of-Day Deviation Risk Score (TOD-RS)

The following procedure was used to derive the Time-of-Day Deviation Risk Score (TOD-RS):

1. **Candidate oscillator selection**: Selected the top 100 oscillatory biomarkers with the largest spline separation (died vs. survived) at 1% FDR, requiring confirmation as oscillatory in the Chronobiome serial study (1% FDR in both datasets).
2. **Reference healthy patterns**: For each candidate biomarker a GAM spline (Eq. 1) was fit in healthy subjects (self-reported good or excellent health) adjusted for age, sex, smoking, Townsend deprivation, fasting time, and storage time to obtain the reference daytime pattern *Ŝ_j_* (*t*).
3. **Per-subject deviation**: For each subject *i* with observation time *t_i_* and z-scaled biomarker value *z_ij_*, the diurnal phase deviation was 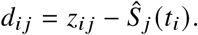
4. **Feature pruning**: Gradient boosted models (GBMs) was applied to select biomarkers predictive of all-cause mortality (training: 70% of proteomics subjects, *n* = 28, 387; test: 30%, *n* = 12, 196)[44]. Sensitivity analysis was performed over varying candidate set sizes(Fig.S11).
5. **Score aggregation**: For subject *i*, 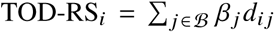, where *β_j_* is the CoxPH coefficient for biomarker *j* and *d_ij_* its time-of-day deviation.

#### Association analyses

A CoxPH model with a penalized spline (pspline) term for standardized TOD-RS, adjusted for age, sex, smoking, Townsend deprivation, fasting time, and storage time, was used to estimate the continuous hazard[55]. For visualization, subjects were stratified into quintiles of TOD-RS and Kaplan–Meier survival curves were plotted[55].

### Artificial Half Cycle Simulations

In the first set of simulations, we generated artificial oscillators using

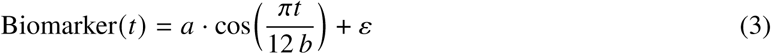

where *t* is the blood draw time (hours since 09:00), *a* is the oscillatory amplitude, and *b* is a frequency-scaling parameter. Amplitudes *a* were sampled uniformly from [−0.3, 0.3] and *b* from [1.0, 1.6]. Non-oscillatory biomarkers were simulated by setting *a* = 0. We generated 3,200 simulated biomarkers (50% null, 50% oscillatory). Noise *ε* was sampled from a normal distribution with mean 0 and standard deviation matched to the empirical variance of each real biomarker category to preserve realistic signal-to-noise characteristics.

Each simulated biomarker was fit with the GAM spline model (Eq. 1) and the distribution of spline F-statistics was compared to that from real data to derive an empirical false discovery threshold.

### Random Permutation Simulations

In the second set of simulations (random permutation), we evaluated whether observed spline term statistics exceeded those expected under a time-independent null by shuffling time-of-day timestamps among subjects for each biomarker. For each of the 3,200 biomarkers, we performed one permutation of its sampling times, refit the GAM model (Eq. 1) with the identical covariate structure, and recorded the spline F-statistic. Permutation destroys any genuine temporal structure while preserving the marginal distribution and covariate relationships, yielding an empirical null distribution.

### Association with ICD-10 and Mortality Outcomes

We tested for the biomarker-outcome association using Cox proportional hazards (CoxPH) survival regression, where events are defined based on the first occurrence of the ICD-10 diagnostic code in the electronic health record (EHR), or the date of death in the case of all-cause mortality. We used the CoxPH method implemented in R survival package[55]. The regressions were adjusted for age, sex, smoking, town deprivation, fasting time, and storage time.

### Replication using Chronobiome Study

To validate the protein oscillators identified in the cross-sectional UKBB data, we used proteomics data from the Chronobiome study[21], which involved serial sampling of 20 healthy individuals over a 48-hour period. For each protein in the Chronobiome dataset, we fitted a generalized additive model (GAM) using the ‘mgcv’ R package. The model included a smoothing spline for time of day (k=6), fixed effects for age group and sex, and a random effect for subject ID to account for the repeated measures. We then compared oscillatory patterns via Pearson correlation on overlapping time-window.

## Data availability

UK Biobank data are available to approved researchers (application procedures at UKBBiobank.ac.uk). Chronobiome data will be made available at www.chronobiome.org.

## Code availability

Code for reproducing the analyses is available at https://github.com/eugenemel/ChronobiomeManuscriptCode

## Acknowledgements

The authors thank colleagues at Calico and the University of Pennsylvania for their support. This research was conducted using UK Biobank Resource application number 18448 and supported by Calico Life Sciences LLC.

## Author contributions

E.M. and G.A.F. conceived the study. E.M. developed theory and methods, performed data analyses, created figures, and wrote the manuscript. A.M., N.F.L. and C.S. contributed to data collection and interpretation. All authors edited, reviewed, and approved the final manuscript.

## Additional information

**Supplementary information** is available for this paper.

**Correspondence and requests for materials** should be addressed to E.M. or G.A.F.

## Supporting Figures

**Figure S1:**
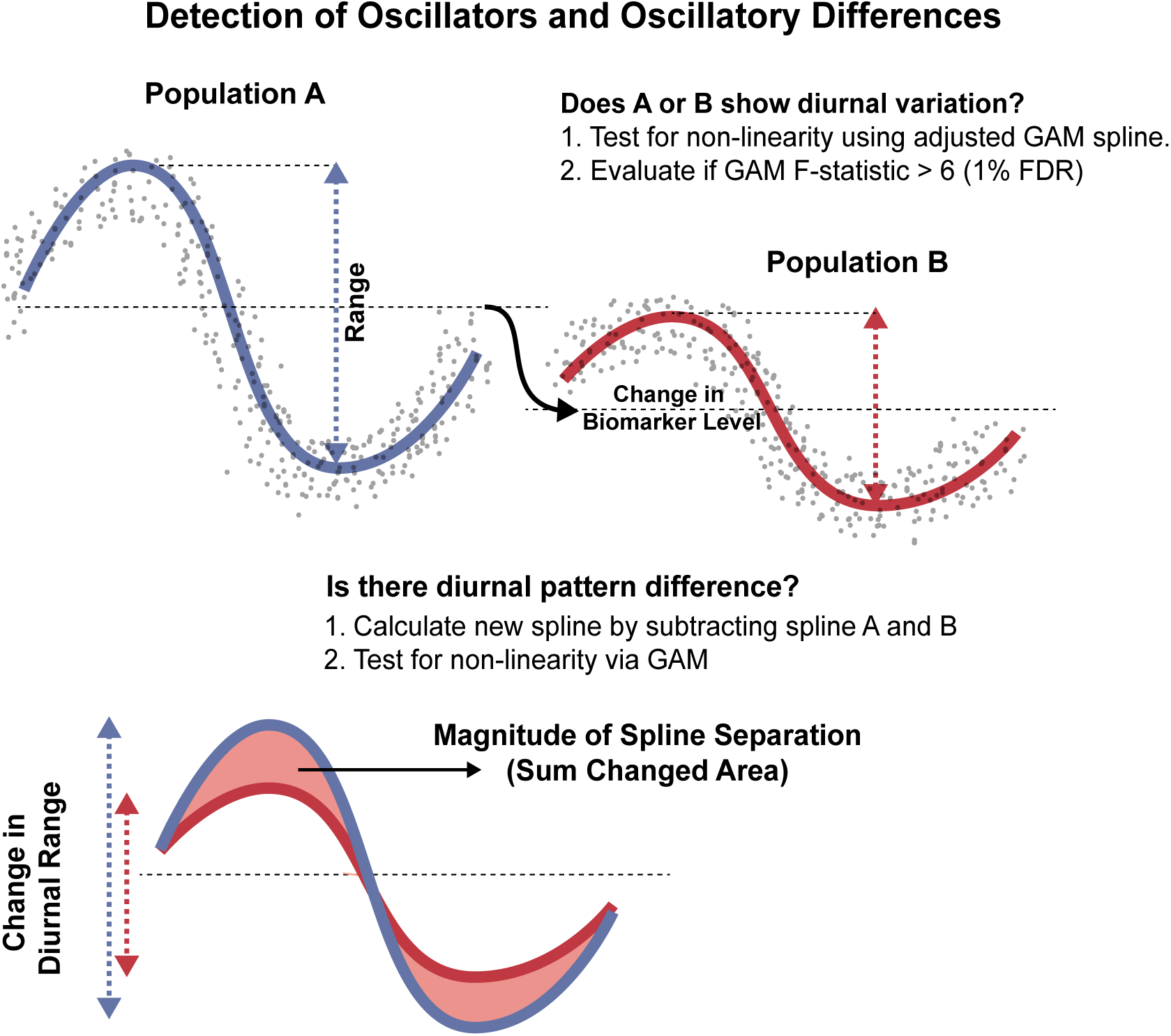
Methods Overview. Detection of oscillatory patterns and their differences using GAM spline regression. Illustration of divergence in spline shapes between two groups. Spline fit of the effect of time of day on biomarker levels adjusted for covariates in groups A and B is shown. Based on simulations and permutation tests, false discovery rate was estimated to be less than 1% at GAM F-statistic>6.0. oscillatory amplitude was defined as the maximum difference between the peak and trough of the spline fit. Assessment of difference in spline shapes was performed by subtracting one spline from another and evaluating non-linearity via GAM regression. Magnitude of spline separation - oscillatory pattern difference - was calculated as the sum of absolute differences between spline shapes.

**Figure S2:**
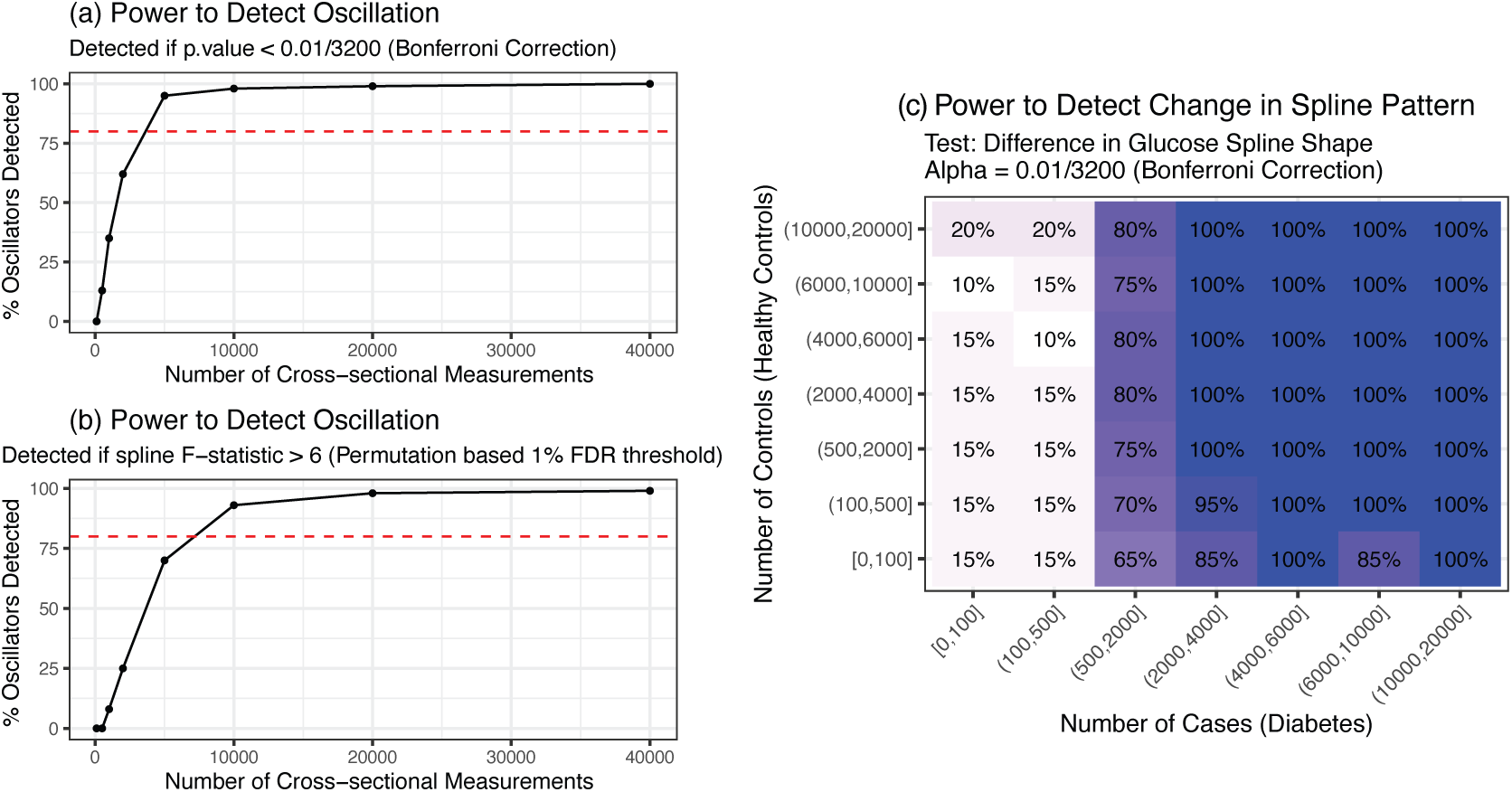
Power Simulation. Power to detect oscillatations in biomarker levels using adjusted GAM spline regression. The simulation is based on sub-sampling measurements from the top 100 oscillatory biomarkers. **a**, Approximately 4,000 cross-sectional measurements are needed to achieve 80% detection power at Bonferroni-corrected *p* = 0.01. **b**, Approximately 7,000 cross-sectional measurements are needed to detect to achieve 80% detection power at 1% FDR (F-statistic> 6). **c**, Power to detect differences in spline shape between two groups of different sizes. Simulation is based on sub-sampling measurements from subjects with and without diabetes using glucose as a known oscillatory biomarker. Greater than 4,000 cases are required to achieve 80% detection power at Bonferroni-corrected *p* = 0.01.

**Figure S3:**
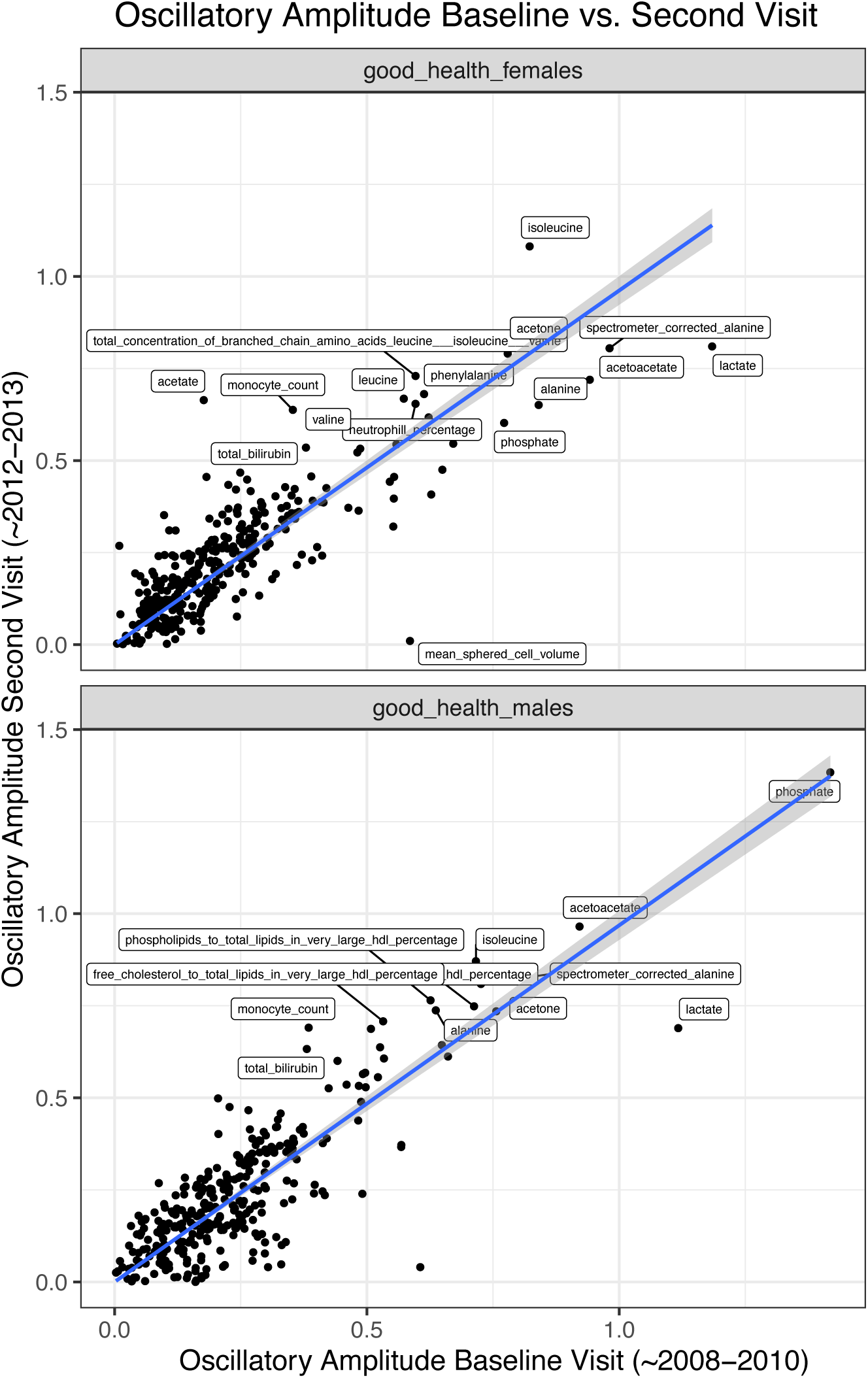
Replication Analysis: Comparison of oscillatory amplitudes for 317 biomarkers in healthy subjects collected at baseline (n∼360,000) and second visit (n∼15,700) in UKBB. oscillatory amplitudes were highly correlated between two visits for both sexes (*R*^2^ ∼ 0.88, *p* < 2 × 10^−16^)

**Figure S4:**
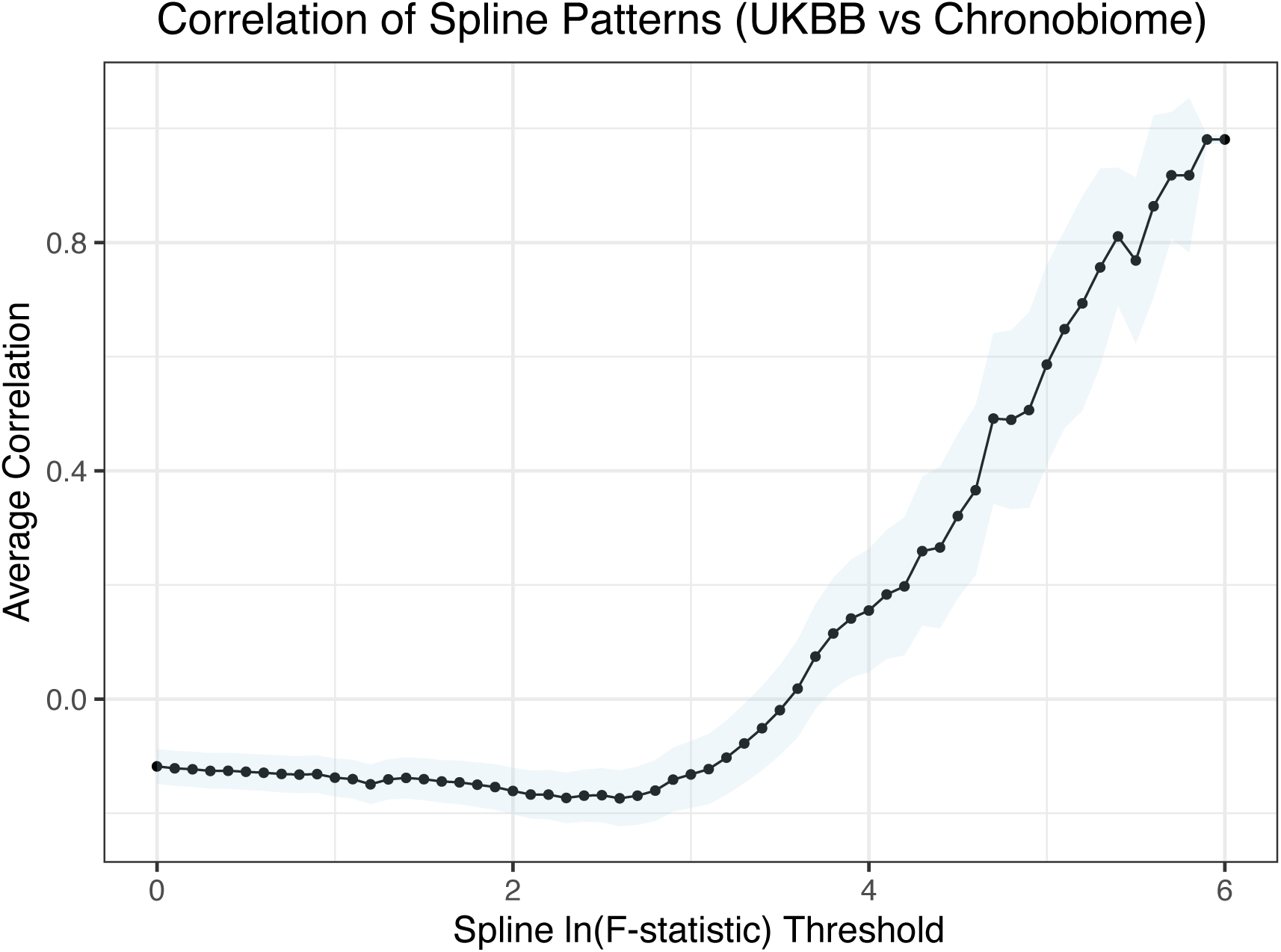
Correlation of oscillatory patterns between serial and cross-sectional measurements. Average correlation of oscillatory pattern between UKBB and serial Chronobiome study as a function of spline ln(F-statistic) cutoff on day-time. Correlation are calculated on spline shape between 9am and 8pm. Strong oscillators (high ln(F-statistic)) are more likely to show high correlation between two studies, with correlations declining for weaker oscillators.

**Figure S5:**
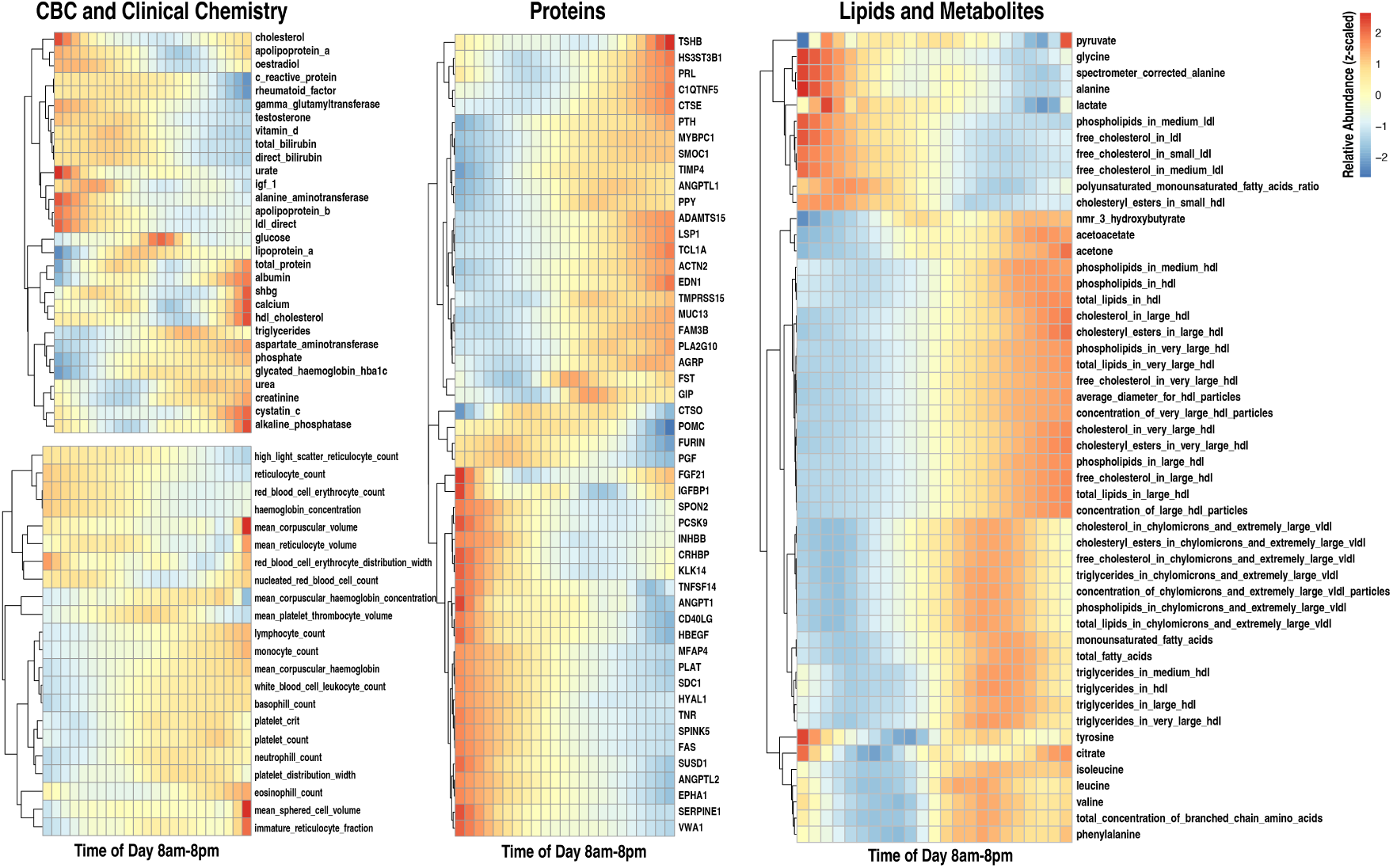
Biomarker Heatmaps. Diurnal phase variation in biomarker levels plotted in half-hour intervals. For proteins and lipids, only the top 50 biomarkers with the largest oscillatory amplitudes are shown. Heatmaps are based on both men and women in healthy subcohort.

**Figure S6:**
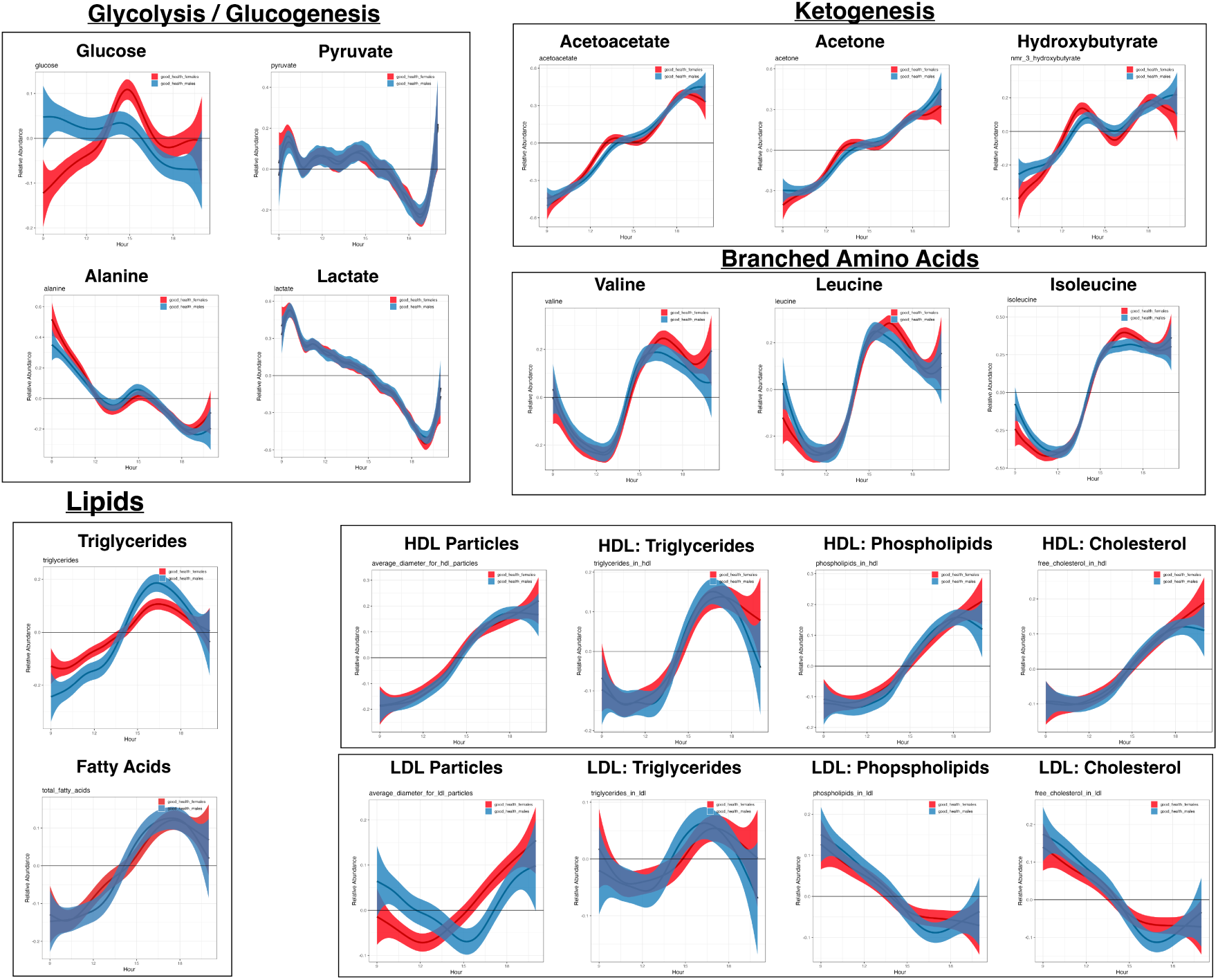
Metabolic and Lipid Oscillators. Diurnal phase variation of key metabolic and lipid intermediates in healthy males (blue) and females (red). Decreasing levels of lactate and pyruvate, increasing levels of ketogenic intermediates, higher levels of triglycerides and fatty acids, and accumulation of phospholipids in HDL and LDL particles, and a sharp transition in essential amino acids in the afternoon were observed.

**Figure S7:**
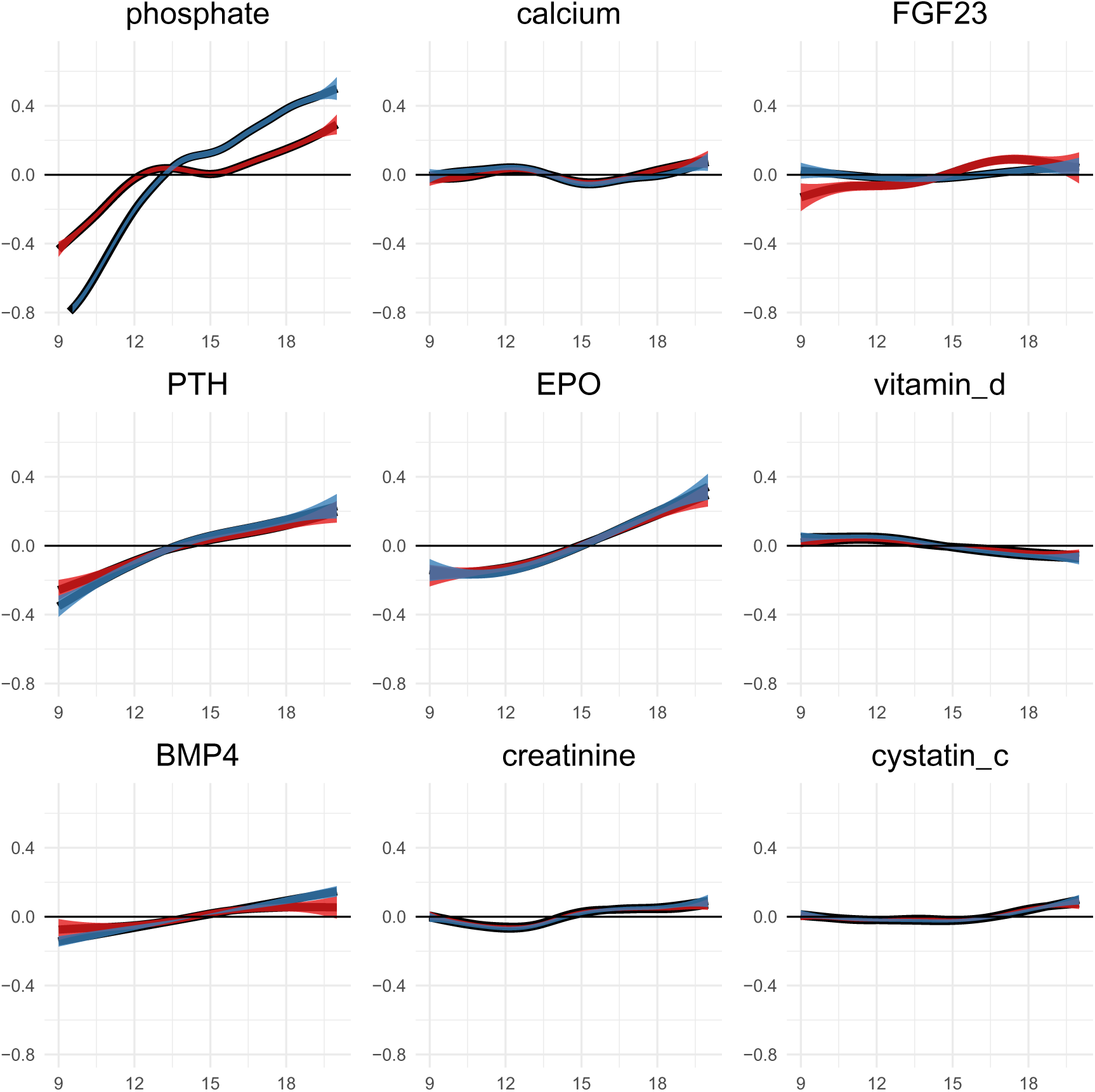
oscillatory patterns in phosphate regulation. Healthy males (blue) vs females (red). Variation in phosphate levels shows a sex-differential effect and is significantly larger than variation in calcium, vitamin D and kidney biomarkers. PTH and EPO show a corresponding increase throughout the day.

**Figure S8:**
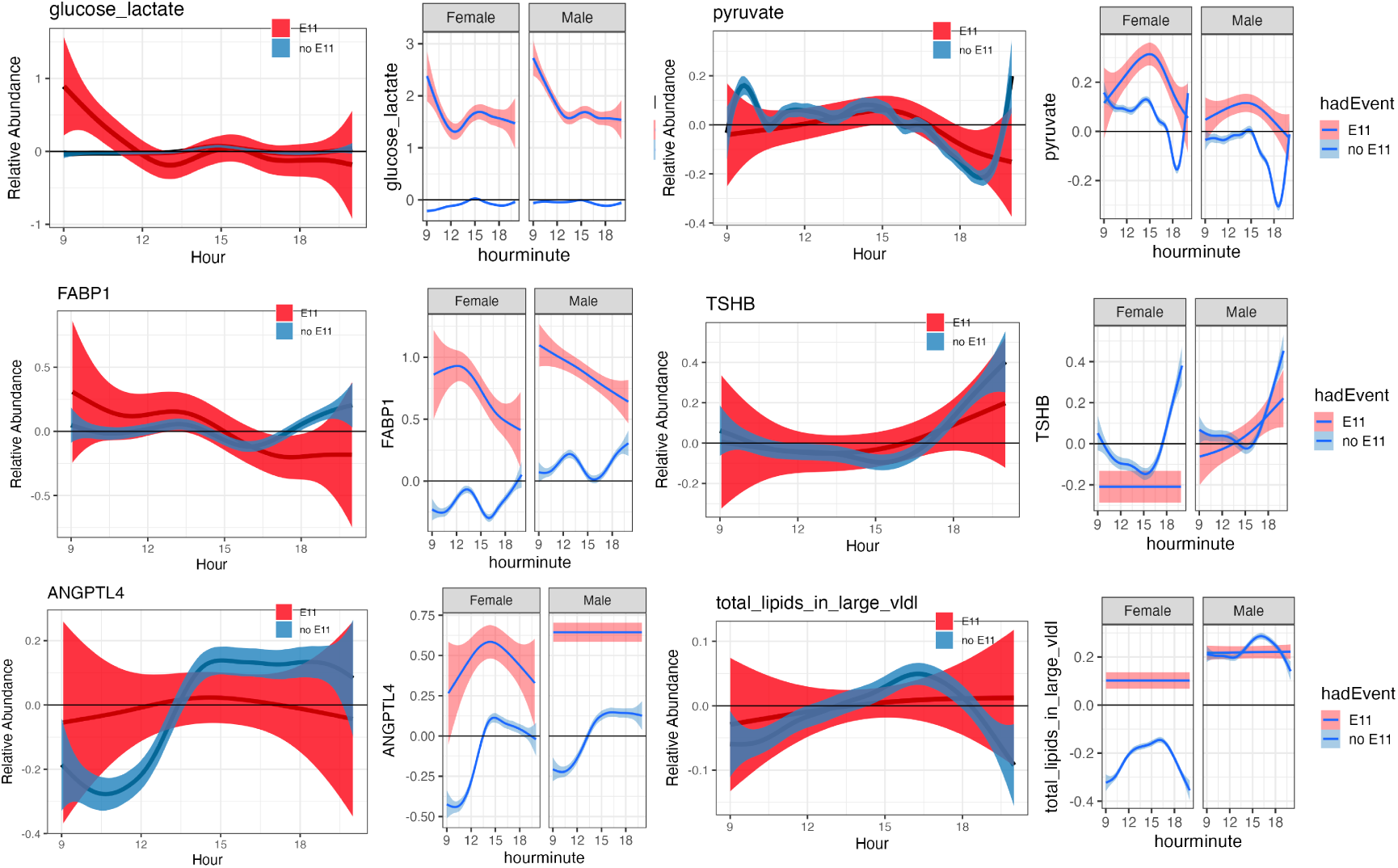
Examples of Pathological Diurnal Phase Variation (Diabetes). Pathological conditions change both level and pattern of oscillations. Adjusted (left panels) and unadjusted sex-stratified spline fits (right panels) are shown for each biomarker.

**Figure S9:**
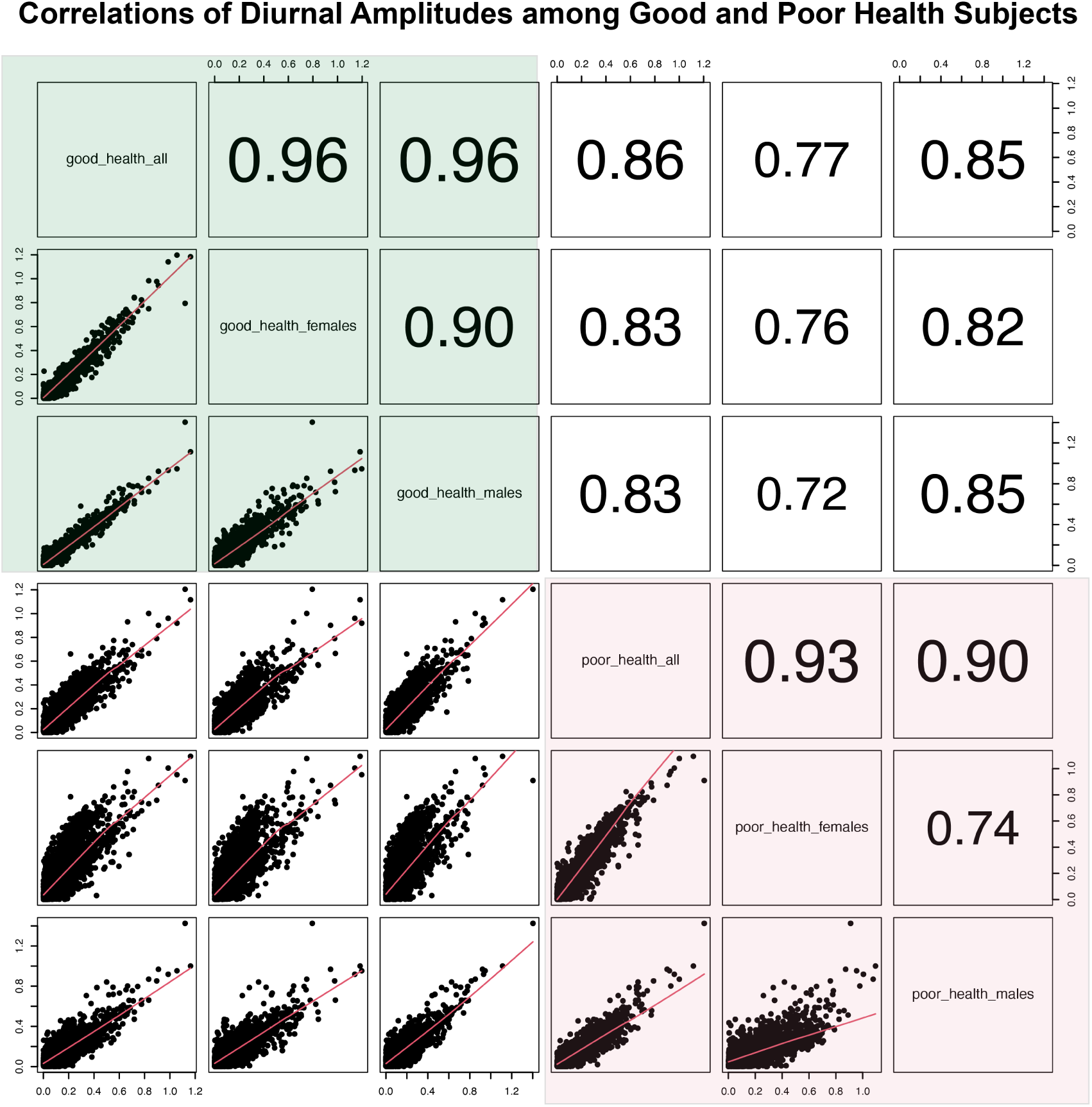
Comparison of oscillatory amplitude in healthy and sick individuals. Correlation of oscillatory amplitudes for subjects stratified by self-reported health rating and sex. Subjects with in good health are shown in light green, while those with poor health are shown in light red. We observed a strong correlation in oscillatory amplitude among healthy subjects (*r* ∼ 0.96) within the healthy subjects(Fig.S9a). There is a significant, but slightly weaker correlation between healthy and sick individuals (*r* ∼ 0.85).

**Figure S10:**
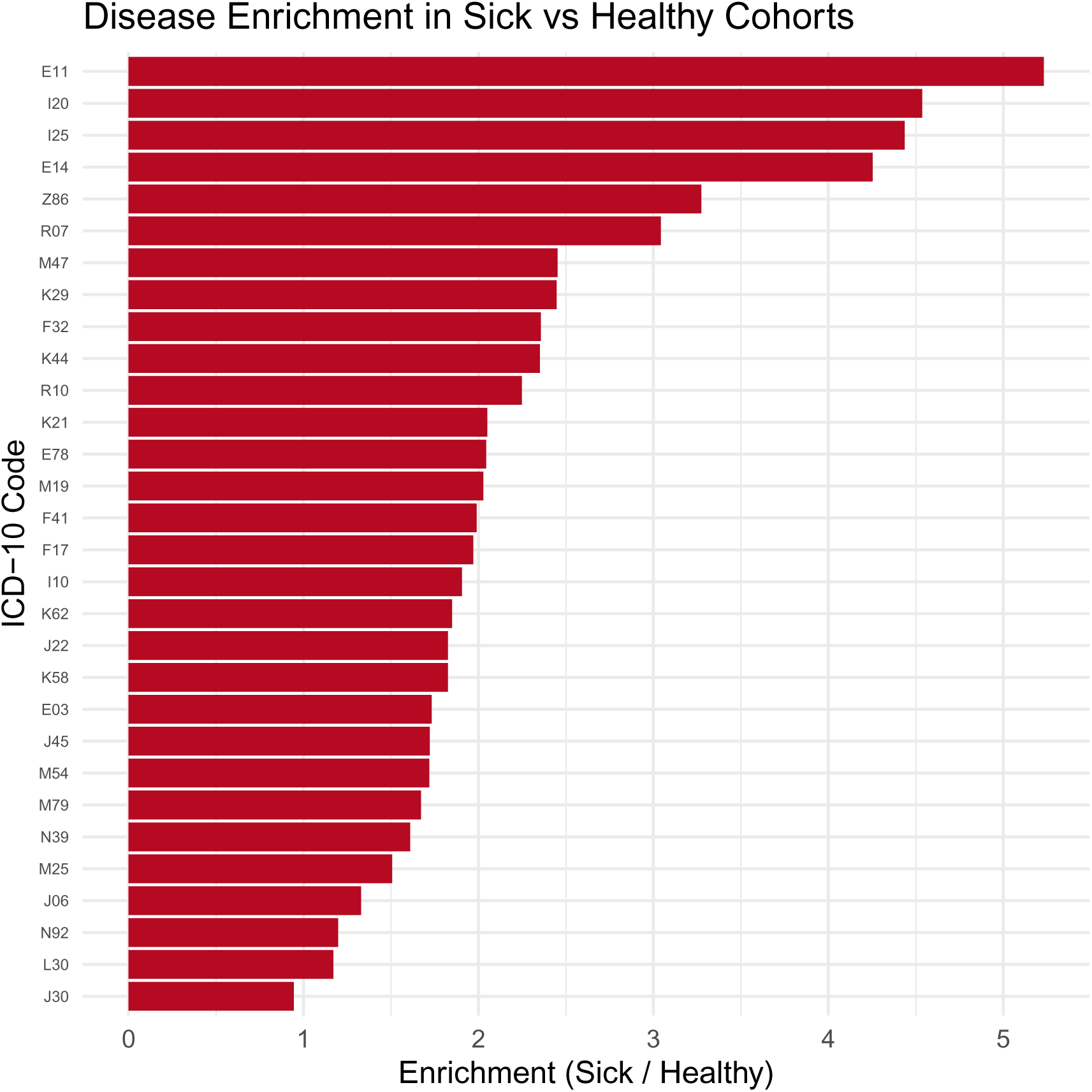
Morbidities enriched among sick subjects. Ratio of incidence rate of ICD-10 codes at baseline between self-reported Fair or Poor health vs those with Good or Excellent healths. Only top 30 conditions are shown. Sick subjects are enriched for cardiovascular, metabolic, respiratory, musculoskeletal health disorders.

**Figure S11:**
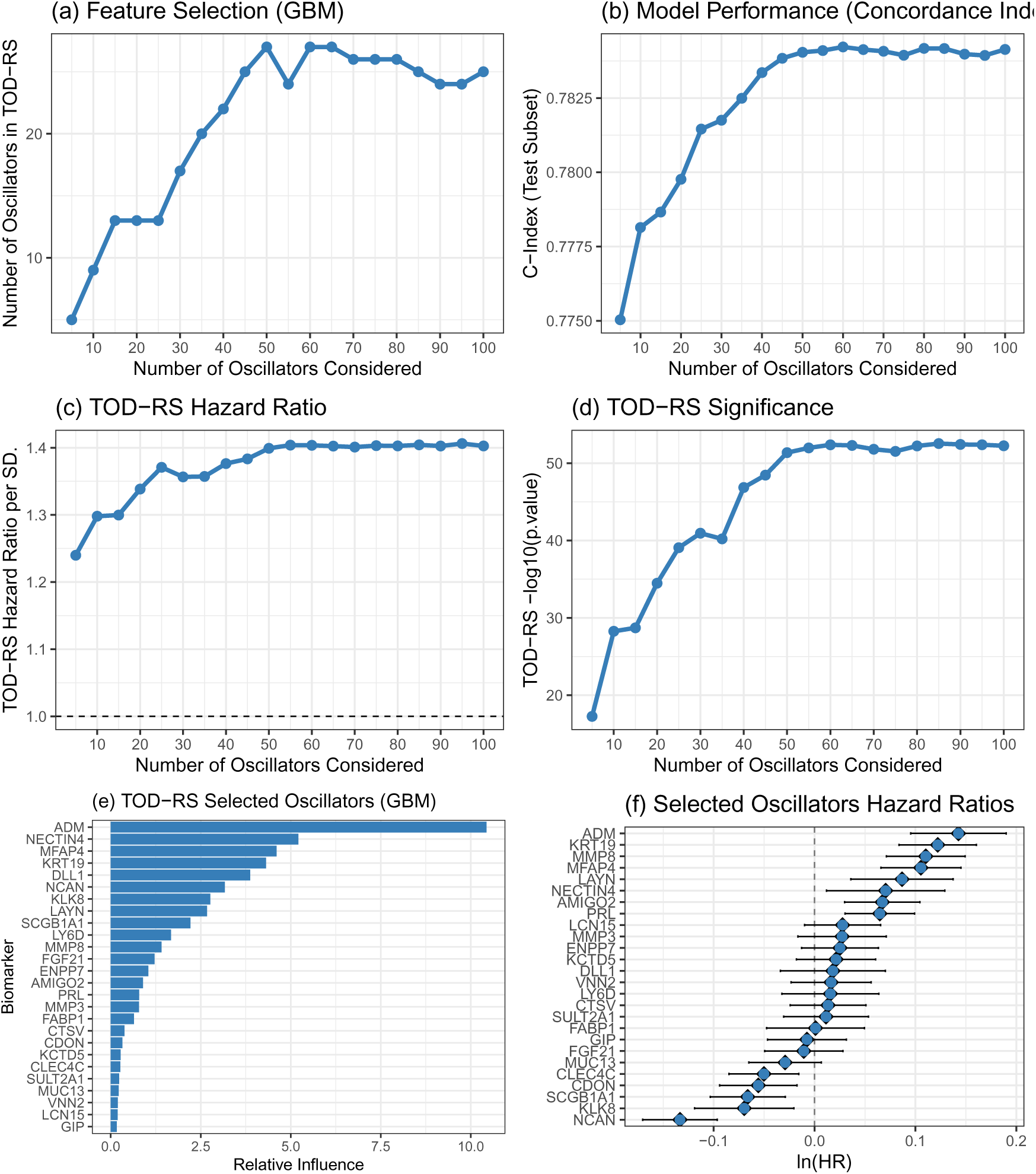
Time-of-Day Deviation Risk Score (TOD-RS) association with all-cause mortality. **a**, Number of selected oscillatory biomarkers retained by GBMs feature pruning vs. candidate set size. **b**, Concordance index (test set) of CoxPH model vs. number of candidate oscillators. **c**, Hazard ratio for TOD-RS (test set) vs. number of candidate oscillators. **d**, − log_10_(*p*) for TOD-RS association. **e**, Relative influence of top oscillators in GBMs. **f**, Individual CoxPH hazard ratios (ln(HR)) for top oscillators.

